# Role of Inflammation in Depressive and Anxiety Disorders, Affect, and Cognition: Genetic and Non-Genetic Findings in the Lifelines Cohort Study

**DOI:** 10.1101/2024.04.17.24305950

**Authors:** Naoise Mac Giollabhui, Chloe Slaney, Gibran Hemani, Éimear M. Foley, Peter J. van der Most, Ilja M. Nolte, Harold Snieder, George Davey Smith, Golam Khandaker, Catharina A. Hartman

## Abstract

**Background:** Low-grade systemic inflammation is implicated in the pathogenesis of various neuropsychiatric conditions affecting mood and cognition. While much of the evidence concerns depression, large-scale population studies of anxiety, affect, and cognitive function are scarce. Importantly, causality remains unclear. We used complementary non-genetic, genetic risk score (GRS), and Mendelian randomization (MR) analyses to examine whether inflammatory markers are associated with affect, depressive and anxiety disorders, and cognitive performance in the Lifelines Cohort; and whether associations are likely to be causal.

**Methods:** Using data from up to 55,098 (59% female) individuals from the Dutch Lifelines cohort, we tested the cross-sectional and longitudinal associations of C-reactive protein (CRP) with (i) depressive and anxiety disorders; (ii) positive and negative affect scores, and (iii) five cognitive measures assessing attention, psychomotor speed, episodic memory, and executive functioning (figural fluency and working memory). Additionally, we examined the association between inflammatory marker GRSs (CRP, interleukin-6 [IL-6], IL-6 receptor [IL-6R and soluble IL-6R (sIL-6R)], glycoprotein acetyls [GlycA]) on these same outcomes (N_max_=57,946), followed by MR analysis examining evidence of causality of CRP on outcomes (N_max_=23,268). In genetic analyses, all GRSs and outcomes were z-transformed.

**Results:** In non-genetic analyses, higher CRP was associated with diagnosis of any depressive disorder, lower positive and higher negative affect scores, and worse performance on tests of figural fluency, attention, and psychomotor speed after adjusting for potential confounders, although the magnitude of these associations was small. In genetic analyses, CRP_GRS_ was associated with any anxiety disorder (β=0.002, *p*=0.037, N=57,047) whereas GlycA_GRS_ was associated with major depressive disorder (β=0.001, *p*=0.036; N=57,047). Both CRP_GRS_ (β=0.006, *p*=0.035, N=57,946) and GlycA_GRS_ (β=0.006, *p*=0.049; N=57,946) were associated with higher negative affect score. Inflammatory marker GRSs were not associated with cognitive performance, except sIL-6R_GRS_ which was associated with poorer memory performance (β=-0.009, *p*=0.018, N=36,783). Further examination of the CRP-anxiety association using MR provided some weak evidence of causality (β=0.12; *p*=0.054).

**Conclusions:** Genetic and non-genetic analyses provide consistent evidence for an association between CRP and negative affect. Genetic analyses suggest that IL-6 signaling could be relevant for memory, and that the association between CRP and anxiety disorders could be causal. These results suggest that dysregulated immune physiology may impact a broad range of trans-diagnostic affective symptoms. However, given the small effect sizes and multiple tests conducted, future studies are required to investigate whether effects are moderated by sub-groups and whether these findings replicate in other cohorts.

## 1. Introduction

Depression affects 300 million individuals worldwide at any given point in time and is the leading cause of mental health-related global disease burden (1–3). Persistent cognitive problems, such as poor memory and concentration, are reported in 11% of adults aged ≥45 years (4) and are frequently observed across a broad range of physical [cancer (35%); COVID-19 (22%); HIV (43%); hepatitis C (50%)] (5–8) and mental health conditions [depression (30%); schizophrenia (50%)] (9, 10). Existing treatments for depression are only modestly effective (11) and almost inexistent for cognitive dysfunction (12, 13). A mechanistic understanding of depression and cognitive dysfunction is urgently needed to inform the development of effective treatments and prevention approaches.

Chronic, low-grade systemic inflammation may represent one such mechanism. Indices of inflammation [e.g., circulating levels of cytokines (e.g., interleukin-6 (IL-6) and acute phase proteins (e.g., C-reactive protein (CRP)] are elevated in individuals with depression compared to controls (14) and inflammatory biomarkers have been linked to specific aspects of depression, such as anhedonia and negative affect (15–17). Further, longitudinal observational studies have found that higher levels of inflammatory biomarkers (e.g., IL-6, CRP) are prospectively associated with higher depressive symptoms (18). Observational studies have linked inflammation with impaired cognition in population-based (19–22) and in physical (5–9, 23–27) and mental health conditions (28–32). Inflammation also impacts neural circuitry relevant to affective disorder and cognitive task performance (33, 34), particularly the hippocampus (35) and striatum (36–39). To date, inflammation-cognition research has primarily relied upon observational data.

Inferring causality from observational studies is a challenge due to confounding (e.g., stress, poor sleep (27)) and reverse causality (i.e., whether inflammation impacts depression/cognition, or vice versa). Mendelian randomization (MR) is a genetic epidemiological method that can test causal relationships by using genetic variants associated with an exposure (e.g., inflammation) as proxies for the exposure (40, 41). As genetic variants are randomly inherited from parents to offspring and are fixed at conception, they are less likely to be associated with confounders and overcome issues of reverse causation (40, 41). Preliminary evidence, using MR, implicate IL-6 and its soluble IL-6 receptor (sIL-6R) in depression (42–45). To date, most MR studies examining the effect of IL-6 on health have focused on circulating IL-6 levels. However, IL-6 signals via multiple pathways (trans-signaling, classical-signaling, and trans-presentation) and there is growing evidence that IL-6 trans-signaling is primarily responsible for the pathogenic inflammatory effects of IL-6 (46, 47). Here, we include variants related to (1) circulating IL-6 levels, and (2) sIL-6R levels (relevant for IL-6 trans-signaling). Causal evidence for CRP and other proinflammatory markers [i.e., Glycoprotein Acetyls (GlycA) a composite biomarker thought to provide a more stable marker of inflammation which reflects the glycosylation of multiple acute-phase proteins (48–51)] on depression are mixed (42, 45, 52–55). Regarding cognition, few studies have examined potential causal relationships with inflammation. MR analyses using available genome-wide association studies (GWAS) report both null results of inflammatory biomarkers on emotion recognition, working memory, response inhibition (56) as well as effects of specific cytokines/chemokines (i.e., Eotaxin, IL-8, MCP1, IL-4) on fluid intelligence (57).

The current study used data from the Lifelines Cohort Study – a large population-based cohort in the Netherlands – to conduct complementary non-genetic and genetic analysis to investigative the causal relationship between inflammation and negative affect, depressive disorders, and cognitive task performance. First, we used cross-sectional and longitudinal non-genetic analysis examine the association between circulating levels of CRP and depression/cognitive performance. Second, we conducted genetic risk score (GRS) and MR analysis to test whether genetic variants regulating levels and activity of CRP, IL-6, and GlycA were causally related with depression/cognitive performance. We also conducted the above analyses on closely related constructs (e.g., anxiety, negative/positive affect), for which associations with inflammation have previously been observed (58–63) but for which considerably less empirical data has been published. We hypothesized that both circulating CRP levels and genetically predicted inflammatory biomarkers (i.e., CRP, IL-6, sIL-6R, and GlycA) would be associated with depression, cognitive task performance, affect, and anxiety.

## 2. Methods and Materials

### 2.1 Participants

Lifelines is a multi-disciplinary prospective population-based cohort study examining in a unique three-generation design the health and health-related behaviors of 167,729 persons living in the North of the Netherlands. It employs a broad range of investigative procedures in assessing the biomedical, socio-demographic, behavioral, physical and psychological factors which contribute to the health and disease of the general population, with a special focus on multi-morbidity and complex genetics (64). This cohort has previously been described in detail (64, 65). In brief, participants were recruited via their general practitioner (49%), participating family members (38%), and self-registration on the Lifelines website (13%). Exclusion criteria for recruitment through the general practitioner included: insufficient knowledge of Dutch language, severe psychiatric or physical illness, limited life expectancy (<5 years). Baseline data included approximately: 140,000 adults (18-65 years), 15,000 children (0-17 years), 12,000 elderly individuals (65+ years). Following baseline assessment, participants are invited to complete an in-person study visit every 5 years (2^nd^ in person follow-up assessment just finished end of 2023). Phenotypic and genotypic data are collected by Lifelines to permit investigation on determinants of health. Data for the current study were drawn from 147,815 individuals who were aged 18+ years at baseline and who did not report a diagnosis that typically impairs cognitive function, specifically Alzheimer’s disease, other dementias, epilepsy, multiple sclerosis, Parkinson’s disease, and stroke. In the non-genetic analyses, the analytic sample is smaller as CRP was assessed in a sub-sample of individuals (N=55,098) as was baseline cognitive performance on the Ruff Figural Fluency Test (N=88,096). The analytic sample is smaller for non-genetic (N≤55,098), GRS (N≤57,946) and MR (N≤23,268) analysis as only a subset of participants who met inclusion criteria had outcome data and (i) CRP data (non-genetic analysis), (ii) genetic data (GRS), or (iii) genetic and CRP data (MR) due to time and cost constraints. Phenotypic data were drawn from both the baseline assessment and the first follow-up assessment; whether a specific measure was assessed at baseline, first follow-up or both assessments is noted for each measure.

### 2.2 Measures

#### 2.2.1 Measures of Cognitive Task Performance

##### Ruff Figural Fluency Test (baseline in-person assessment)

The Ruff Figural Fluency Test (RFFT) is a reliable and valid measure of figural fluency, a dimension of executive function (66), measured at the baseline assessment. It is likely that performance on the RFFT also depends on other cognitive abilities, such as processing speed (67, 68). Participants were asked to draw as many unique designs as possible within 60 seconds by connecting dots in different patterns. The task is composed of five parts, with each part containing 35 identical five-dot patterns (with or without distractors). The total number of unique designs was used as the dependent variable in the analyses, consistent with previous studies (69, 70). In Lifelines, the RFFT was administered to all participants until April 2012, and subsequently in a random half of the sample. Data from participants who failed to generate a single unique design per trial (*n* = 181) were deemed invalid and removed.

##### Cogstate Test Battery (first follow-up in-person assessment)

Assessments included in the Cogstate Test Battery consisted of four tasks and took approximately 10-15 minutes to complete. Each task was designed to tap into specific cognitive domains: detection task (psychomotor speed), identification task (attention), one-back task (working memory), and one card learning task (episodic memory), although, like in the RFFT, processing speed likely plays a role in all tasks. For each task, outcomes recommended by Cogstate were selected, specifically: log10 transformed response time in millisecond (detection and identification tasks) and arcsine-transformed response accuracy (one-back and one card learning tasks). For the detection and identification tasks, higher values reflect poorer performance and for the one-back and one card learning tasks, higher values equal better performance. Data cleaning involved excluding participants with a high number of errors. The percentage of successful trials per Cogstate task was high, averaging 66% (*n* = 85,050; *SD* = .11) on the episodic memory task, 91% (*n* =85,053; *SD* = .17) on the visual attention task, 92% (*n* =85,053; *SD* = .20) on the psychomotor speed task, and 90% (*n* = 85,051; *SD* = .15) on the working memory task. A small number of participants exhibiting implausibly low accuracy rates indicative of poor effort, failure to comprehend task instructions, or technical errors were excluded from analyses. Specifically individuals with an accuracy rate less than: 25% on the episodic memory task (*n* = 231), 40% on the visual attention task (*n* = 2,878), 45% on the psychomotor speed task (*n* = 3,914), and 35% on the working memory task (*n* = 1,330). For more details on the Cogstate Test Battery, see Supplementary Methods 1.1.

#### 2.2.2 Clinical Assessments

##### Anxiety and Depressive Disorders (baseline and first follow-up in-person assessments)

The Mini International Neuropsychiatric Interview – Simplified (MINI) is a reliable, valid, and brief structured interview that was designed to screen for psychiatric disorders (71–73). Lifelines used an adapted version of a Dutch translation of the MINI that was administered by trained interviewers at baseline and self-administered on location at follow-up – details on the version used in Lifelines have previously been published (74). Participants were considered to meet criteria for any depressive disorder if they met the Diagnostic and Statistical Manual of Mental Disorders (DSM)-IV criteria for Major Depressive Disorder (MDD) *or* dysthymia at the time of the interview. Impairment was assessed in the MINI for dysthymia but not depression and consequently, impairment was not used as a criterion for MDD. Any anxiety disorder refers to meeting current criteria for any one of the following conditions that was assessed using the MINI: panic disorder, agoraphobia, social phobia, or Generalized Anxiety Disorder (GAD). We used four diagnostic groups as outcome variables: MDD, any depressive disorder, GAD, and any anxiety disorder.

##### Positive and Negative Affect Schedule (baseline in-person assessment)

The Positive and Negative Affect Schedule (PANAS) is composed of two subscales which are designed to assess positive and negative affect (75). Each subscale has 10 items (examples of items include ‘excited’ on positive subscale; ‘upset’ on negative subscale). Participants are asked to rate the extent that they experienced each item during the last four weeks on a five-point scale (ranging from ‘not at all’ to ‘extremely’). The outcome is the summed score on each subscale, which ranges from 10 to 50 (higher value reflects higher positive or negative affect, respectively). The PANAS has been shown to be reliable and valid (76).

#### 2.2.3 C-reactive Protein (baseline in-person assessment)

Participants gave blood samples before 10AM via venipuncture following an overnight fast. Complete details on blood specimen data collection have previously been reported (64, 70). Due to assay costs, CRP was assessed in approximately 35% of Lifelines participants and data were available for 55,098 individuals in the analytic sample. CRP was quantified using three separate methods over the course of baseline assessment (Method 1: 12.90% of total CRP values assessed in serum; CardioPhase hsCRP; Method 2: 84.58% of total CRP values, assessed in plasma; CardioPhase high sensitivity (hs)CRP, Siemens Healthcare Diagnostics, Marburg, Germany; Method 3: 2.52% of total CRP values; assessed in plasma; CRPL3, Roche Diagnostics, Mannheim, Germany). Assay methods 2 and 3 were identical and only differed in terms of the manufacturer. A conversion formula (new = 0.92 x old - 0.01) was derived from an internal validation using 39 samples, according to the AMC (alternative method comparison, Deming Regression) protocol in order that Method 1 could be compared with Method 2 and 3 (70). For CardioPhase hsCRP, the intra-assay coefficient of variability was 3.45% and the inter-assay coefficient of variability was 3.15%. For CRPL3, the intra-assay coefficient of variability was 4.15% and the inter-assay coefficient of variability was 5.8%.

#### 2.2.4 Genetic Data

Genotype data were available for a subgroup of participants in Lifelines. Genotyping was conducted using three chip arrays: (i) Illumina CytoSNP-12 Bead Chip v2 array (N=17,033), (ii) Infinium Global Screening Array (GSA) Beadchip-24 v1.0 (N=38,030), (iii) FinnGen Thermo Fisher Axiom ® custom array (Affymetrix; N=29,166). For details on quality checks (QC’s) and imputation conducted by Lifelines, see Supplementary Methods 1.2. Following Lifelines QC’s, the total sample size for participants who met criteria for this study: CytoSNP (N=14,942), GSA (N=31,810) and Affymetrix (N=26,334). We applied additional QC’s which included removing: (i) one of the duplicates (individuals who were genotyped on more than one chip) and first-degree relatives between chips, (ii) non-European individuals (identified by Lifelines), and (iii) genetic outliers (identified by Lifelines); see Supplementary Figure 1 for more details. This resulted in a total of 58,713 participants with genetics data included in this study (CytoSNP N=7,632; GSA N=24,975; Affymetrix N=26,106). For more details on the genetic data in Lifelines, see Supplementary Methods 1.2.

#### 2.2.5 Covariates

Covariates included age, sex, educational attainment, body mass index (BMI) and health status. Age, sex, and educational attainment were self-reported by participants. Educational attainment was determined using a single-item question and was categorized by Lifelines as: low [no education, primary education, lower/preparatory vocational education, lower general secondary education (leaving secondary school aged >16 years)], moderate (intermediate vocational education/apprenticeship, higher secondary education), and high (higher vocational education, university). We recoded educational attainment so that higher values represent lower educational attainment. To estimate body mass index (BMI), height was measured to the closest 0.1 cm and body weight was measured without shoes to 0.1 kg precision. For health status, a composite measure was created counting several self-reported chronic medical conditions related to increased levels of inflammation (i.e., arthritis, asthma, coeliac disease, Crohn’s disease, diabetes, and psoriasis); we then categorized participants into those with no relevant chronic medical condition, 1, 2 or 3+ conditions.

### 2.3 Analyses

Analyses were conducted in R version 4.1.1.(77)

#### 2.3.1 Non-genetic Analyses

Multivariable linear and logistic regression models were estimated using base functions in R (i.e., ‘lm’, ‘glm’). CRP was transformed by natural log to impose a normal distribution.

#### 2.3.2 Genetic Risk Scores

Genetic risk scores (GRS) were calculated to determine whether GRS for inflammatory markers (CRP, IL-6, IL-6R, sIL-6R, GlycA) were associated with depression/anxiety, affect and cognitive outcomes. To create GRS for each inflammatory marker, we identified genetic variants (single nucleotide polymorphisms [SNPs]) associated with these proteins in large available GWAS or using SNP lists from previous publications, see Table 1. For details on the GWAS used and accessing summary statistics, please see Supplementary Methods 1.3 and Supplementary Tables 1-2. The following criteria were used to identify SNPs from GWAS for each inflammatory marker: (i) *p*-value < 5 x 10^-8^, (ii) linkage disequilibrium clumping (r^2^=0.01, kb=1000 based on the European-clustering individuals in the 1000 genomes reference panel) using *ld_clump()* (78) in the *ieugwasr* package, (iii) minor allele frequency > 0.01. In the primary analysis, we restricted the SNP set to *cis* variants (SNPs +/- 1Mb from protein coding gene based on Genome Reference Consortium Human Build used in the GWAS) (79–84). The reason for restricting to *cis* variants in the primary analysis is because, due to their proximity to the protein coding gene, they are more likely to be valid instruments, as they are more likely to influence mRNA expression and protein levels (thus being less pleiotropic) (85). For GlycA, which does not have a single coding gene due to its composite nature, we used the largest available GWAS in our primary analysis. In our secondary analyses, we used both *cis* and *trans* variants from GWAS (i.e., we did not restrict to *cis* variants). Each SNP list (Table 1) was used to create a weighted GRS for each Lifelines participant. Specifically, the risk alleles were weighted by the effect size (beta) reported in the GWAS/previous study and then summed to provide a risk score. Any SNP identified in GWAS/previous study that was not available in Lifelines was replaced with a proxy (where possible) that had *r*^2^ > 0.8 (using *LDproxy_batch* function in EUR population in *R*), rsID (SNP name) available, SNP available in full summary statistic GWAS, and in Lifelines (86, 87). GRS were created in Plink v1.90 (88) and continuous phenotypes were standardized within each chip (z-scored) for direct comparison (CRP levels were log transformed but not standardized).

**Table 1.** Large available genome-wide association studies and previous publications used to identify single nucleotide polymorphisms associated with systemic inflammatory markers.

| **Marker** | **Author of GWAS/**<br>**Instrument** | **Sample**<br>**size** | **Outcome** | **SNPs**<br>**Identified** | **SNPs**<br>**available on**<br>**each chip** |
| --- | --- | --- | --- | --- | --- |
| **Primary analysis** |  |  |  |  |  |
| CRP | (Said et al., 2022) | 575,531 | Circulating CRP | 19 | 16 |
| IL-6R | (Ahluwalia et al., 2021) | 52,654 | Circulating IL-6 | 2 | 2 |
|  | (Sarwar et al., 2012) | 27,185 | Circulating IL-6 | 1 | 1 |
| sIL6R | (Rosa et al., 2019) | 3,301 | Circulating sIL6R | 34 | 34 |
| GlycA | (Borges, 2020) | 115,078 | Circulating GlycA | 87 | 73-74 |
| **Secondary analysis** |  |  |  |  |  |
| CRP | (Said et al., 2022) | 575,531 | Circulating CRP | 485 | 445-446 |
| IL-6 | (Ahluwalia et al., 2021) | 52,654 | Circulating IL-6 | 3 | 2-3 |

CRP = C-reactive protein; IL-6 = interleukin-6, IL-6R = IL-6 receptor, sIL6R = soluble IL-6R

To adjust for relatedness within each chip, two approaches were taken. The primary approach applied the GRAMMAR method (89) and the secondary approach involved re-running analyses removing close relatives (up to first-degree, up to second-degree, and up to third-degree), see Supplementary Materials 1.4 and 1.5 for details on how we identified close relatives. We then ran regression models predicting each outcome using the standardized GRS, including top 10 genetic PCs (calculated on merged Lifelines genotype data), age, sex, and chip. Maximum sample size for analyses: no relatives within chips removed (N=58,713), up to first-degree removed (N=50,955), up to second-degree removed (N=50,255), up to third-degree removed (N=48,880). Unadjusted analyses are also reported in the Supplementary Tables 5 and 6 for comparison.

#### 2.3.3 Mendelian randomization

To conduct MR using individual level data and two-stage least squares regression, genetic data, exposure data, and outcome data are required. As only CRP is available within the Lifelines cohort (IL-6 and GlycA are not currently available), only this inflammatory marker could be assessed in the MR analysis. Where there was evidence of associations between CRP GRS and outcomes, we followed this up with MR to assess potential causality. Three key assumptions are necessary for valid inferences from MR: (i) genetic variants are robustly associated with the exposure, (ii) genetic variants are not associated with potential confounders, (iii) genetic variants are associated with the outcome only via the exposure. Two-stage least squares regressions were conducted using the *AER* package (90). Analyses were GRAMMAR adjusted for relatedness and all regression models adjusted for age, sex, and chip. To check MR assumptions, we ran linear regressions to test whether CRP GRS were associated with circulating CRP levels in participants with both genetic and CRP data available (*n* = 23,607) using the GRAMMAR method. We also checked whether any inflammatory marker GRS (CRP, IL-6, IL-6R, sIL-6R, GlycA) were associated with potential confounders (BMI, current smoking status, educational attainment; all models were adjusted for age, sex, and chip).

## 3. Results

The characteristics of the Lifelines cohort sample are described in Table 2 and Pearson correlations between study variables are presented in Table 3.

**Table 2.** Lifelines Cohort Sample Characteristics at Baseline and First Follow-up Assessment.

| <u>Measures</u> | <u>Cohort Analyses</u><br><u>(n = 147,815)</u> | <u>Genetic Analyses</u><br><u>(n = 58,713)</u> |
| --- | --- | --- |
| **Baseline Assessment** |  |  |
| Age [Mean (*SD*)] | 44.52 (13.12)<br>n = 147,815 | 43.04 (13.56)<br>n = 58,713 |
| Sex (% Female) | 59 %<br>n = 147,815 | 60%<br>n = 58,695 |
| Education (N %) | n = 146,050 | n = 58,112 |
| - Lower | 43,750 (30%) | 16,359 |
| - Moderate | 57,785 (40%) | 23,770 |
| - Higher | 44,515 (30%) | 17,983 |
| Body mass index | 26.05 (4.34)<br>n = 147,719 | 25.81 (4.27)<br>n = 58,680 |
| RFFT (Unique designs) [Mean (*SD*)] | 81.50 (22.94)<br>n = 88,096 | 82.46 (23.01)<br>n = 36,563 |
| Any Depressive Disorder (Current Major Depression or<br>Dysthymia) | 3.4%<br>n = 141,045 | 2.9%<br>n = 56,861 |
| Major Depressive Episode (Current) | 2.1%<br>n = 141,538 | 1.8%<br>n = 57,048 |
| Any Anxiety Disorder (panic disorder, agoraphobia, social<br>phobia, or GAD ) | 7.8%<br>n = 141,538 | 7.2%<br>n = 57,048 |
| Generalized Anxiety Disorder | 4.2%<br>n = 141,539 | 3.8%<br>n = 57,048 |
| Negative Affect Score [Mean (*SD*)] | 20.71 (13.12) | 20.70 (5.22) |
|  | n = 139,217 | n = 57,964 |
| Positive Affect Score [Mean (SD)] | 35.37 (4.25)<br>n = 139,217 | 35.37 (4.19)<br>n = 57,964 |
| C-reactive protein level (mg/L), [Median (IQR),<br>Mean (SD)] | 1.2 (.60, 2.80),<br>2.61 (4.76)<br>n = 55,098 | 1.2 (2.2)<br>2.62 (4.60)<br>n = 23,607 |
| **First follow up** |  |  |
| Any Depressive Disorder (Current Major Depression or<br>Dysthymia) | 4.1%<br>n = 77,758 | Not used |
| Major Depressive Episode (Current) | 3.0%<br>n = 77,758 | Not used |
| Any Anxiety Disorder (panic disorder, agoraphobia, social<br>phobia, or GAD ) | 8.3%<br>n = 77,758 | Not used |
| Generalized Anxiety Disorder | 5.9%<br>n = 77,758 | Not used |
| Cogstate: Episodic Memory (Accuracy), Mean (SD) | 0.96 (0.12)<br>n = 84,819 | 0.96 (0.12)<br>n = 36,798 |
| Cogstate: Working Memory (Accuracy), Mean (SD) | 1.31 (0.19)<br>n = 83,721 | 1.32 (0.19)<br>n = 36,363 |
| Cogstate: Visual Attention (Response Time), Mean (SD) | 2.69 (0.09)<br>n = 82,175 | 2.68 (0.09)<br>n = 35,743 |
| Cogstate: Psychomotor Speed (Response Time), Mean (SD) | 2.56 (0.16)<br>n = 81,139 | 2.55 (0.16)<br>n = 35,314 |
| Lower = no education, primary education, lower/preparatory vocational education, lower general secondary education; Moderate = intermediate vocational education/apprenticeship, higher secondary education; Higher = higher vocational education, university; IQR = Inter Quartile Range. |  |  |

**Table 3.** Bivariate Correlations of Study Variables for 147,815 Participants.

| Measure | 2. | 3. | 4. | 5. | 6. | 7. | 8. | 9. | 10. | 11. | 12. | 13. | 14. | 15. | 16. | 17. | 18. | 19. | 20. | 21. |
| --- | --- | --- | --- | --- | --- | --- | --- | --- | --- | --- | --- | --- | --- | --- | --- | --- | --- | --- | --- | --- |
| 1. T1 CRP (log-transformed) | -0.03 | -0.03 | 0.04 | 0.03 | -0.07 | 0.04 | 0.03 | 0.03 | -0.05 | 0.03 | 0.14 | 0.11 | 0.36 | 0.10 | 0.03 | 0.03 | 0.03 | 0.03 | 0.02 | 0.03 |
| 2. T2 Episodic Memory<sup>B</sup> | - | 0.31 | -0.16 | -0.16 | 0.21 | -0.02 | -0.03 | -0.05 | -0.02 | -0.19 | 0.01 | 0.22 | -0.08 | -0.04 | -0.02 | -0.01 | -0.03 | -0.03 | -0.01 | -0.01 |
| 3. T2 Working Memory<sup>B</sup> |  | - | -0.17 | -0.17 | 0.20 | -0.02 | -0.02 | -0.04 | .00<sup>a</sup> | -0.20 | -.01<sup>a</sup> | -0.2 | -0.07 | -0.04 | -0.02 | 0 | -0.03 | -0.02 | -0.01 | -0.01 |
| 4. T2 Psychomotor Speed<sup>C</sup> |  |  | - | 0.63 | -0.30 | .01<sup>a</sup> | .00<sup>a</sup> | 0.04 | .00<sup>a</sup> | 0.39 | .00<sup>a</sup> | 0.21 | 0.08 | 0.04 | 0.01 | -0.02 | 0.01 | 0 | -0.01 | -0.02 |
| 5. T2 Visual Attention<sup>C</sup> |  |  |  | - | -0.34 | 0.01 | .00<sup>a</sup> | 0.05 | -0.02 | 0.43 | -0.01 | 0.20 | 0.09 | 0.05 | 0.02 | -0.01 | 0.02 | 0.01 | 0.01 | -0.01 |
| 6. T1 RFFT |  |  |  |  | - | -0.04 | -0.02 | -0.07 | -0.07 | -0.32 | 0.03 | -0.35 | -0.11 | -0.08 | -0.04 | -0.01 | -0.04 | -0.02 | -0.02 | 0.01 |
| 7. T1 Depression |  |  |  |  |  | - | 0.25 | 0.27 | -0.2 | -0.02 | 0.05 | 0.07 | 0.04 | 0.07 | 0.35 | 0.19 | 0.8 | 0.21 | 0.40 | 0.18 |
| 8. T2 Depression |  |  |  |  |  |  | - | 0.21 | -0.13 | -0.05 | 0.03 | 0.05 | 0.03 | 0.05 | 0.20 | 0.41 | 0.22 | 0.83 | 0.21 | 0.42 |
| 9. T1 Negative Affect |  |  |  |  |  |  |  | - | -0.21 | -0.05 | 0.17 | 0.08 | -0.01 | 0.06 | 0.32 | 0.26 | 0.25 | 0.17 | 0.30 | 0.22 |
| 10. T1 Positive Affect |  |  |  |  |  |  |  |  | - | -0.02 | -0.01 | -0.1 | -0.02 | -0.02 | -0.17 | -0.13 | -0.18 | -0.1 | -0.18 | -0.11 |
| 11. T1 Age |  |  |  |  |  |  |  |  |  | - | -0.04 | 0.23 | 0.19 | 0.09 | -0.01 | -0.06 | -0.01 | -0.04 | -0.04 | -0.06 |
| 12. T1 Female |  |  |  |  |  |  |  |  |  |  | - | 0.02 | -0.06 | 0.03 | 0.07 | 0.07 | 0.04 | 0.03 | 0.05 | 0.06 |
| 13. T1 Education |  |  |  |  |  |  |  |  |  |  |  | - | 0.17 | 0.08 | 0.06 | 0.03 | 0.07 | 0.05 | 0.04 | 0.02 |
| 14. T1 Body mass index |  |  |  |  |  |  |  |  |  |  |  |  | - | 0.13 | 0.03 | 0.01 | 0.04 | 0.03 | 0.02 | 0.01 |
| 15. T1 Health Status |  |  |  |  |  |  |  |  |  |  |  |  |  | - | 0.07 | 0.04 | 0.06 | 0.05 | 0.05 | 0.04 |
| 16. T1 ANX |  |  |  |  |  |  |  |  |  |  |  |  |  |  | - | 0.28 | 0.31 | 0.17 | 0.72 | 0.2 |
| 17. T2 ANX |  |  |  |  |  |  |  |  |  |  |  |  |  |  |  | - | 0.17 | 0.37 | 0.22 | 0.84 |
| 18. T1 MDE |  |  |  |  |  |  |  |  |  |  |  |  |  |  |  |  | - | 0.20 | 0.38 | 0.16 |
| 19. T2 MDE |  |  |  |  |  |  |  |  |  |  |  |  |  |  |  |  |  | - | 0.17 | 0.40 |
| 20. T1 GAD |  |  |  |  |  |  |  |  |  |  |  |  |  |  |  |  |  |  | - | 0.21 |
| 21. T2 GAD |  |  |  |  |  |  |  |  |  |  |  |  |  |  |  |  |  |  |  | - |

Probability <sup>a</sup> = $P > .05$ ; <sup>B</sup> = higher values equal better performance; <sup>C</sup> = higher values equal poorer performance; T1 = Time 1 (Baseline); T2 = Time 2 (First Follow-up); RFFT = Ruff Figural Fluency Test; Health Status. = Number of Medical Conditions Reported; for values $\leq .001$ and $\geq -.001$ , values were rounded to 0.

### 3.1 Association of CRP with affect, depressive and anxiety disorders, and cognitive task performance

The association of (log-transformed) CRP with: (i) clinical outcomes (i.e., MDD, any depressive disorder, GAD, any anxiety disorder), (ii) positive and negative affect, and (iii) five cognitive measures [RFFT (executive functioning), detection task (psychomotor speed), identification task (attention), one-back task (working memory), and one card learning task (episodic memory)] are illustrated in Table 4, both unadjusted and adjusted for covariates.

**Table 4.**
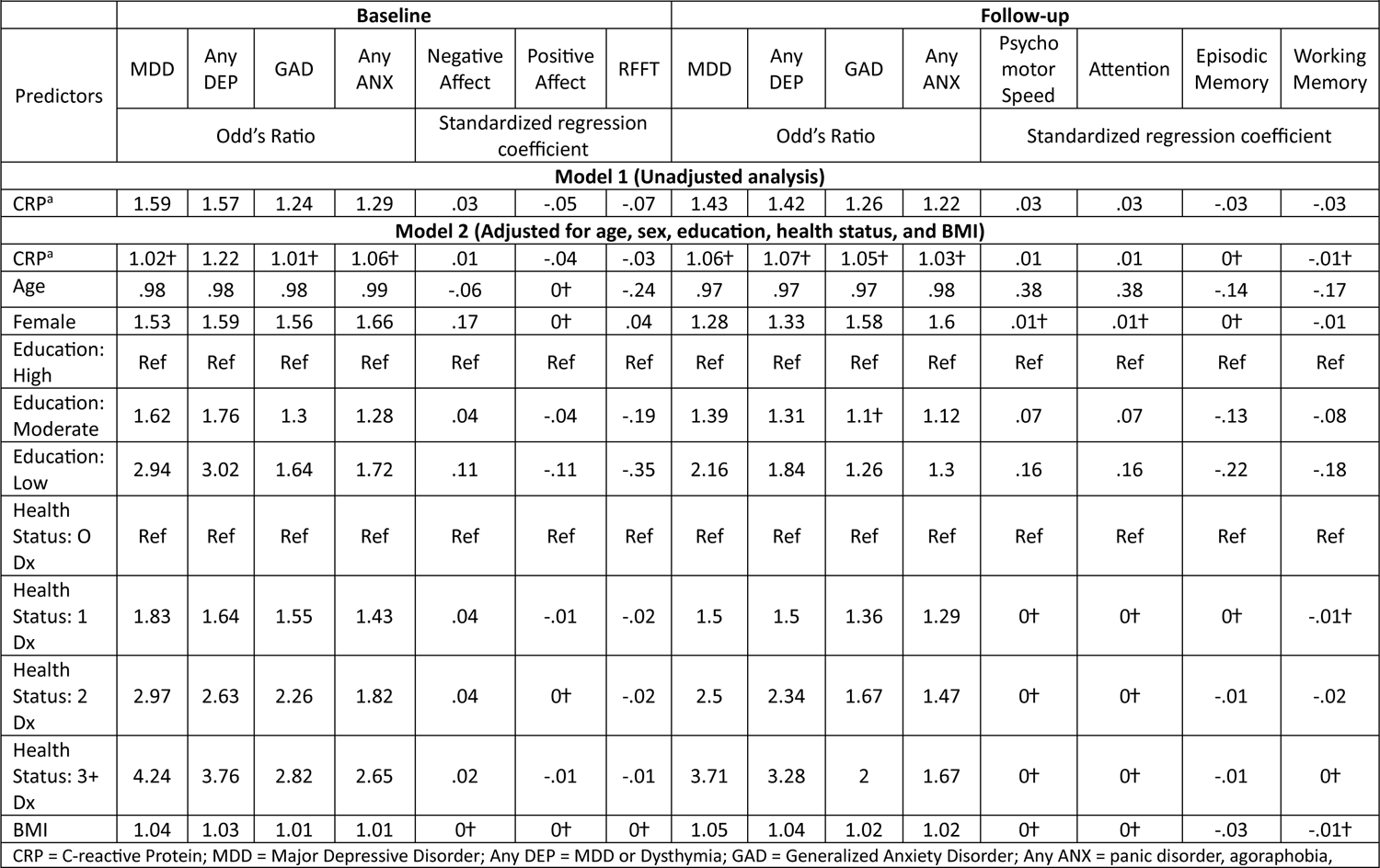
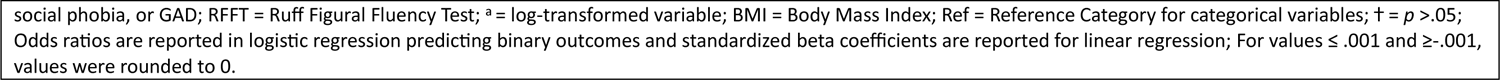
Associations of CRP levels with affect, depressive and anxiety disorders, and cognitive task performance in the Lifelines cohort Please note: point estimates do not include 95% confidence intervals (and N, p-value) as we do not currently have access to the Lifelines Cohort Workspace, the Cohort will allow us access to data for response to reviewer comments, we will add these (p-value, 95% CI, N) as necessary during review.

|  | Baseline |  |  |  |  |  |  | Follow-up |  |  |  |  |  |  |  |
| --- | --- | --- | --- | --- | --- | --- | --- | --- | --- | --- | --- | --- | --- | --- | --- |
| Predictors | MDD | Any DEP | GAD | Any ANX | Negative Affect | Positive Affect | RFFT | MDD | Any DEP | GAD | Any ANX | Psycho motor Speed | Attention | Episodic Memory | Working Memory |
|  | Odd's Ratio |  |  |  | Standardized regression coefficient |  |  | Odd's Ratio |  |  |  | Standardized regression coefficient |  |  |  |
| Model 1 (Unadjusted analysis) |  |  |  |  |  |  |  |  |  |  |  |  |  |  |  |
| CRP<sup>a</sup> | 1.59 | 1.57 | 1.24 | 1.29 | .03 | -.05 | -.07 | 1.43 | 1.42 | 1.26 | 1.22 | .03 | .03 | -.03 | -.03 |
| Model 2 (Adjusted for age, sex, education, health status, and BMI) |  |  |  |  |  |  |  |  |  |  |  |  |  |  |  |
| CRP<sup>a</sup> | 1.02† | 1.22 | 1.01† | 1.06† | .01 | -.04 | -.03 | 1.06† | 1.07† | 1.05† | 1.03† | .01 | .01 | 0† | -.01† |
| Age | .98 | .98 | .98 | .99 | -.06 | 0† | -.24 | .97 | .97 | .97 | .98 | .38 | .38 | -.14 | -.17 |
| Female | 1.53 | 1.59 | 1.56 | 1.66 | .17 | 0† | .04 | 1.28 | 1.33 | 1.58 | 1.6 | .01† | .01† | 0† | -.01 |
| Education: High | Ref | Ref | Ref | Ref | Ref | Ref | Ref | Ref | Ref | Ref | Ref | Ref | Ref | Ref | Ref |
| Education: Moderate | 1.62 | 1.76 | 1.3 | 1.28 | .04 | -.04 | -.19 | 1.39 | 1.31 | 1.1† | 1.12 | .07 | .07 | -.13 | -.08 |
| Education: Low | 2.94 | 3.02 | 1.64 | 1.72 | .11 | -.11 | -.35 | 2.16 | 1.84 | 1.26 | 1.3 | .16 | .16 | -.22 | -.18 |
| Health Status: 0 Dx | Ref | Ref | Ref | Ref | Ref | Ref | Ref | Ref | Ref | Ref | Ref | Ref | Ref | Ref | Ref |
| Health Status: 1 Dx | 1.83 | 1.64 | 1.55 | 1.43 | .04 | -.01 | -.02 | 1.5 | 1.5 | 1.36 | 1.29 | 0† | 0† | 0† | -.01† |
| Health Status: 2 Dx | 2.97 | 2.63 | 2.26 | 1.82 | .04 | 0† | -.02 | 2.5 | 2.34 | 1.67 | 1.47 | 0† | 0† | -.01 | -.02 |
| Health Status: 3+ Dx | 4.24 | 3.76 | 2.82 | 2.65 | .02 | -.01 | -.01 | 3.71 | 3.28 | 2 | 1.67 | 0† | 0† | -.01 | 0† |
| BMI | 1.04 | 1.03 | 1.01 | 1.01 | 0† | 0† | 0† | 1.05 | 1.04 | 1.02 | 1.02 | 0† | 0† | -.03 | -.01† |

CRP = C-reactive Protein; MDD = Major Depressive Disorder; Any DEP = MDD or Dysthymia; GAD = Generalized Anxiety Disorder; Any ANX = panic disorder, agoraphobia,
social phobia, or GAD; RFFT = Ruff Figural Fluency Test; <sup>a</sup> = log-transformed variable; BMI = Body Mass Index; Ref = Reference Category for categorical variables; † = $p > .05$ ; Odds ratios are reported in logistic regression predicting binary outcomes and standardized beta coefficients are reported for linear regression; For values $\leq .001$ and $\geq -.001$ , values were rounded to 0.

Notably, CRP was associated with a greater likelihood of meeting criteria for a range of clinical outcomes, with a numerically greater likelihood consistently reported for depression as compared to anxiety at baseline and first follow-up assessment. However, these associations were attenuated after controlling for confounding by age, sex, education, health status, and BMI. Higher CRP was also associated with higher negative affect, lower positive affect, and worse cognitive task performance, although the magnitude of associations was generally very small and negligible after controlling for covariates.

### 3.2 Associations of GRSs for inflammatory markers with affect, depressive and anxiety disorders

In the primary analysis, CRP_GRS_ (*cis*) was associated with a higher negative affect score (beta: 0.006; 95% CI: 0.0005 to 0.012, *p*=0.035, N=57,946) and increased risk of any anxiety disorder (beta: 0.002, 95% CI: 0.0001 to 0.004, *p*=0.037, N=57,047). GlycA_GRS_ was associated with higher negative affect score (beta: 0.006, 95% CI: 0.00002 to 0.012, *p*=0.049; N=57,946) and increased risk of MDD (beta: 0.001, 95% CI: 0.0001 to 0.002; *p*=0.036; N=57,047). Other inflammatory marker GRSs were not associated with depressive/anxiety disorders or affect scores (*p*s ≥ 0.15). In the secondary analysis, there was evidence that CRP_GRS_ (*genome-wide*) was associated with increased risk of any anxiety disorders (beta: 0.002, 95% CI: 0.0003 to 0.004, *p*=0.023, N=57,047). There was little evidence that other inflammatory marker GRSs were associated with depressive/anxiety disorders or affect (*p*s ≥ 0.16). For all results, see Figure 1 and Supplementary Table 4. All sensitivity analyses removing differing degrees of related individuals (up to 1^st^-degree, up to 2^nd^-degree, up to 3^rd^-degree) within chips (non-GRAMMAR method) did not substantially alter results, see Supplementary Tables 7-12. Although, there was slightly stronger evidence for associations between GlycA_GRS_ and negative affect score (*p*s ≤ 0.015) and between CRP_GRS_ (*cis*) and negative affect score (*p*s ≤ 0.033).

**Figure 1.**
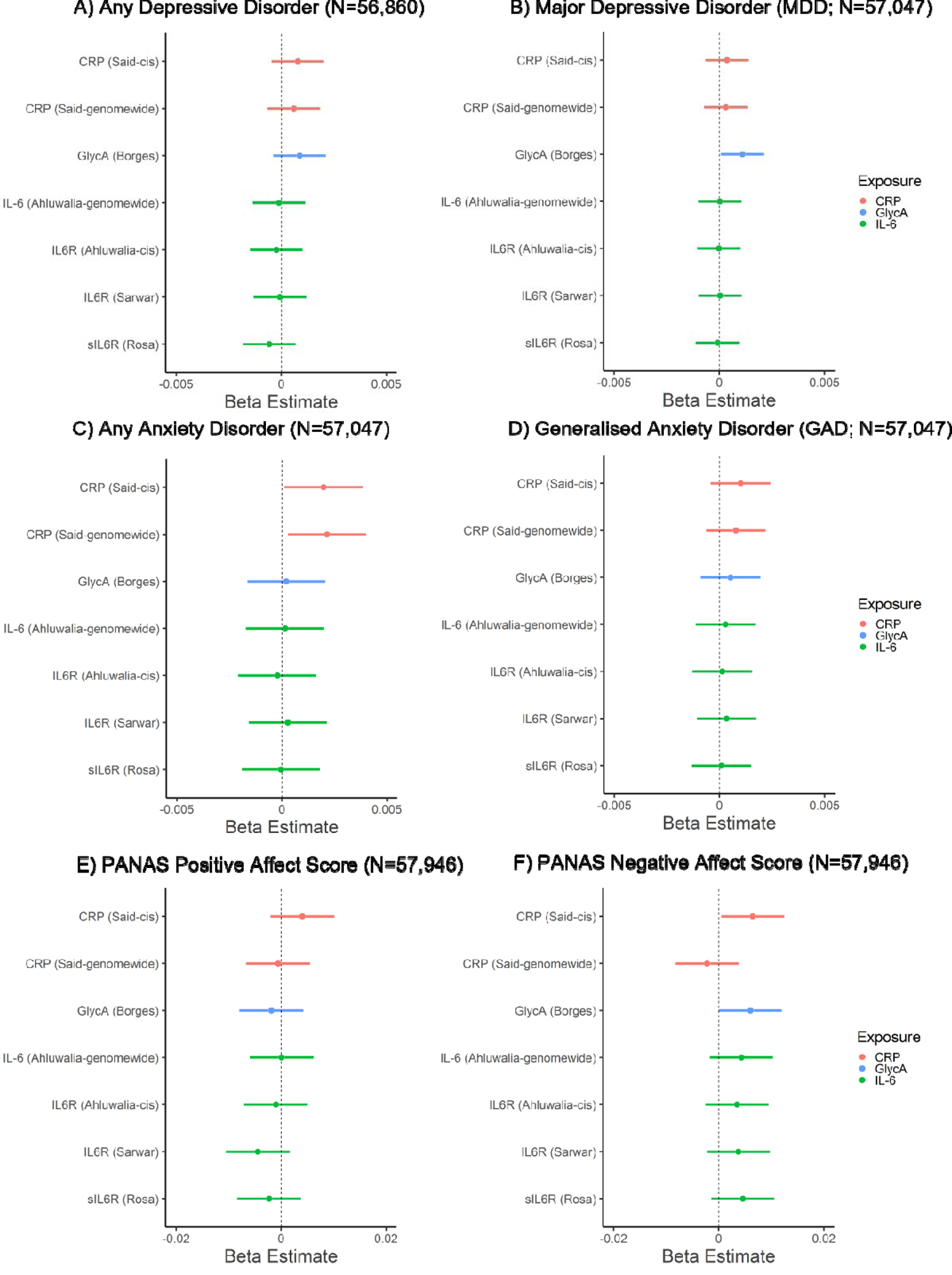
Associations of genetic risk scores for inflammatory markers with mood, anxiety disorders and positive and negative affect scores.

### 3.3 Association of GRS for inflammatory markers and cognitive task performance

In primary analyses, inflammatory marker GRSs were not associated with performance on cognitive tasks (*p*s ≥ 0.14), except for sIL-6R_GRS_ which was negatively associated with episodic memory performance (one card learning task accuracy; beta: −0.009, 95% CI: −0.017 to −0.002, *p*=0.018, N=36,783), see Figure 2 and Supplementary Table 4. In secondary analyses, inflammatory markers GRSs (*genome-wide*) were not associated with performance on cognitive tasks (*p*s ≥ 0.22). For all results, see Figure 2 and Supplementary Table 4. Sensitivity analyses after removing related individuals within chips (non-GRAMMAR method) did not alter the results, see Supplementary Tables 10-12.

**Figure 2.**
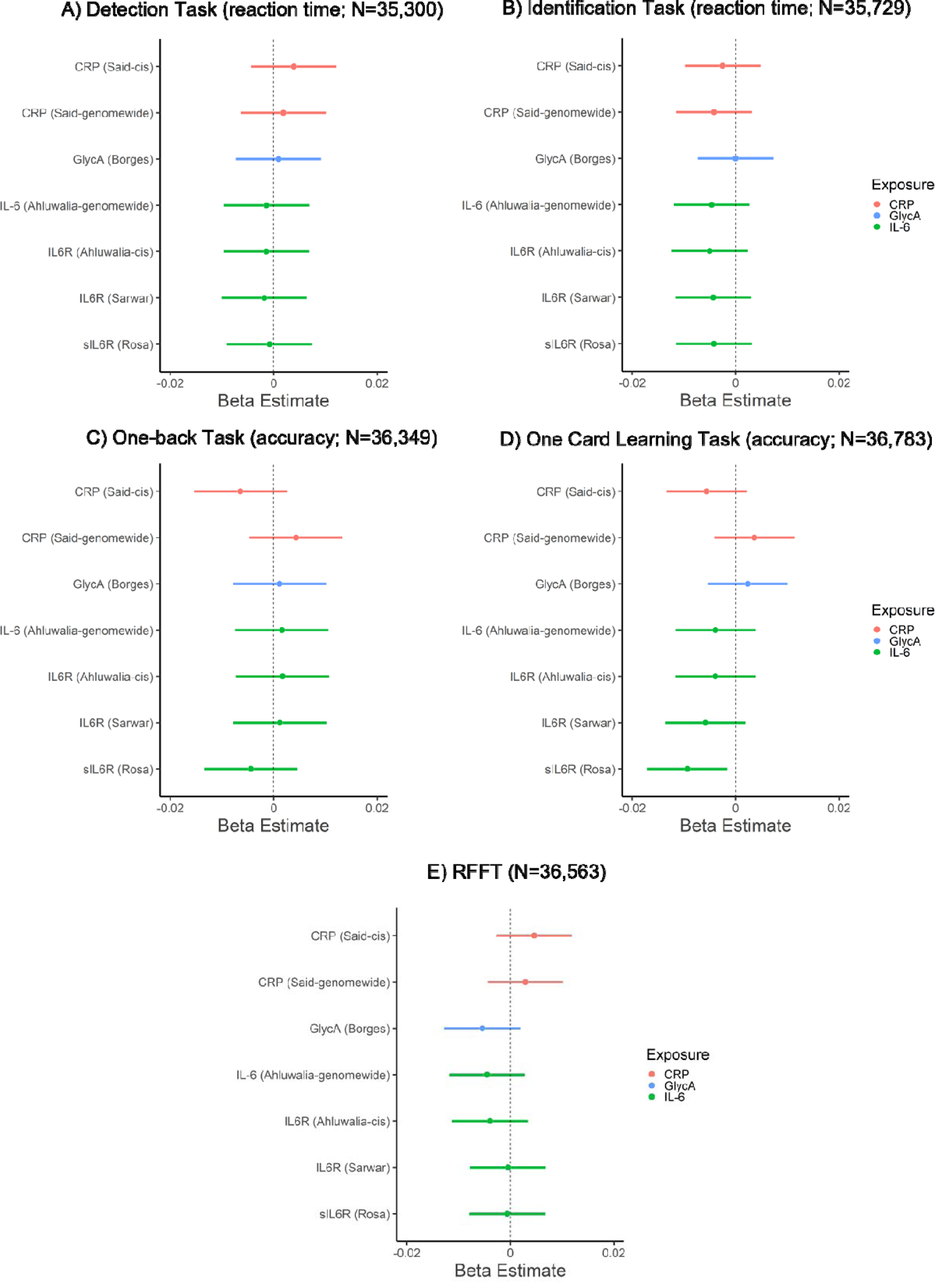
Associations of genetic risk scores for inflammatory markers with cognitive task performance.

### 3.4 Testing potential causality between CRP, negative affect, and anxiety disorders using Mendelian randomization with individual level data

CRP genetic instruments had F-statistics > 10 (158 for *cis* GRS, 1045 for *genome-wide* GRS), indicating adequate instrument strength (91). For tests on the MR assumptions, see Supplementary Results 2.2. There was weak evidence that genetically-proxied CRP (*cis*) causally increased the risk of any anxiety disorders (beta: 0.12, *p*=0.054, N=22,154), and little evidence on negative affect (beta: 0.27, *p*=0.16; N=23,268). Sensitivity analysis removing related individuals did not alter overall conclusions. The overall pattern of results for the non-genetic and genetic analyses are visualized in Figure 3.

**Figure 3.**
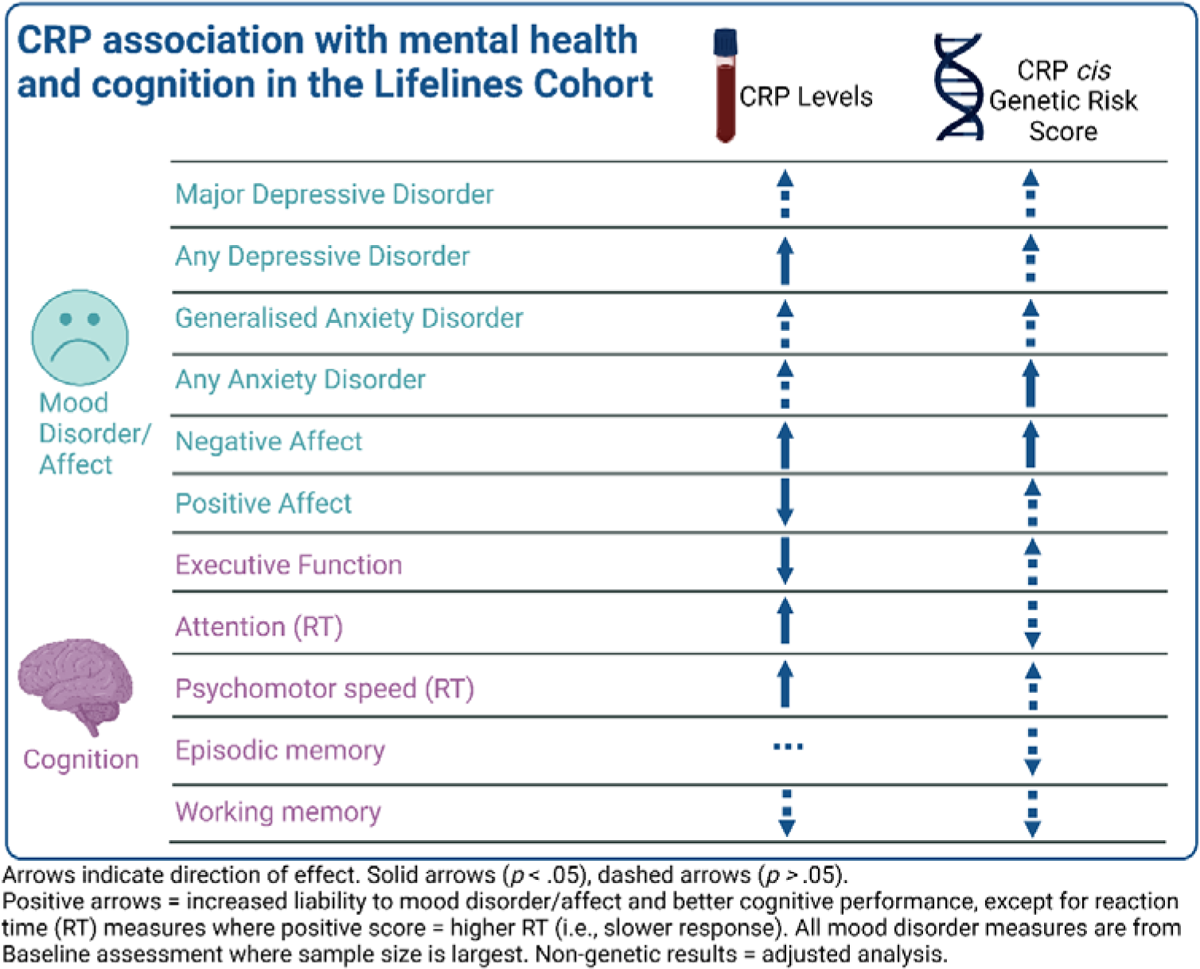
Visualisation of the overall pattern of results for CRP in the cohort and genetic analyses.

## 4. Discussion

We conducted complementary non-genetic and genetic analyses to interrogate the relationship between inflammatory markers and affect, depressive and anxiety disorders, and cognitive task performance using data from the Lifelines cohort. In non-genetic analyses, higher CRP was associated with diagnosis of any depressive disorder, positive and negative affect scores, figural fluency, attention, and psychomotor speed after adjusting for potential confounders, although the magnitude of these associations was generally small. In genetic analyses, genetic risk scores for CRP (CRP_GRS_) and GlycA_GRS_ were both associated with higher negative affect score. CRP_GRS_ was associated with any anxiety disorder whereas GlycA_GRS_ was associated with major depressive disorder. Inflammatory marker GRSs were not associated with cognitive task performance, except for soluble IL-6R_GRS_ which was associated with poorer memory performance. Individual level MR provided weak evidence for a causal effect of CRP on any anxiety disorder. Genetic and non-genetic analyses provided consistent evidence for an association, albeit small, of CRP on negative affect. Genetic analyses suggest that IL-6 signaling could be relevant for memory, and that the association between CRP and anxiety disorders could be potentially causal.

### 4.1 Affect

Prior studies have generally found inflammation to be associated with higher levels of negative affect and lower levels of positive affect, although findings are primarily based on medical populations (92–94) and small community samples (15, 95, 96). To our knowledge, this is the first large, population-based study to find small but consistent associations of higher CRP with higher negative affect and lower positive affect, both unadjusted and adjusted for age, sex, education, health status and BMI. Interestingly, both CRP and GlycA genetic risk scores were associated with higher levels of negative affect, but not positive affect. This consistent association across non-genetic and genetic analyses may reflect the effect of inflammation on a range of emotional states beyond the cardinal features of depression (i.e., sadness/anhedonia), which aligns with prior research linking inflammation with fear and irritability (63, 97). Prior work has shown that inflammation is differentially associated with a specific clinical presentation characterized by anhedonia and somatic/neurovegetative symptoms (e.g., fatigue, altered sleep and appetite changes) and further work is needed that more accurately characterize an inflammatory phenotype in depression (98, 99).

### 4.2 Depression

These data add to a growing body of evidence evaluating the role of inflammation as measured by circulating CRP levels in the etiology of depression. The results of non-genetic analyses broadly aligns with results from the UK Biobank cohort in terms of (i) prevalence estimates of depression and anxiety, (ii) robust univariate associations between CRP and depression and anxiety, which were generally no longer statistically significant when controlling for covariates, and (iii) stronger univariate associations for CRP and depression when compared to anxiety (45). It has long been noted that variables being conceptualized as confounds that require statistical adjustment (e.g., BMI, medical illness) may, in fact, be key mechanisms in the pathophysiology of inflammatory depression (100, 101). As such, attenuation of associations following adjustment for covariates would not, by itself, indicate a non-causal relationship. Indeed, inflammation may increase risk for depression via increasing the risk of inflammation-related physical multimorbidity (e.g., cardiovascular disease) (42) – a hypothesis that requires further investigation.

In genetic analyses, there was also little evidence of an association between CRP_GRS_ and depression outcomes, although there was evidence suggesting GlycA_GRS_ increases liability to MDD. The null CRP findings are consistent with previous MR studies showing no evidence of effect in MDD (54, 55, 102). However, the MR literature of CRP on depression is mixed with some studies reporting CRP to decrease (45) or increase (42) risk for depression. It is unclear what accounts for these mixed findings, but potential factors may include CRP SNP selection, definition and/or measurement of depression, statistical power, and selection bias (see Supplementary Discussion). In contrast, MR studies have shown more consistent findings for the potential causal role of IL-6 on depression (43–45, 103). This is similar to MR findings for coronary heart disease, where IL-6 but not CRP have been shown to play a potential causal role (104, 105). Consequently, studies on a broader range of immune markers (e.g. cytokines, immune cells) and specific immune pathways would be more useful to understand the role of inflammation in depression, rather than CRP which is a non-specific marker of systemic immune activation (106).

### 4.3 Cognitive Task Performance

We observed relatively small effects of CRP on cognitive task performance, and in genetic analysis only sIL-6R_GRS_ was associated with poor memory performance out of all inflammatory markers and cognitive tasks examined. Our findings contribute to inconsistent findings across population-based cohorts assessing circulating inflammatory biomarkers and cognitive performance where associations observed in population-based studies (22, 30, 107) often are not large in magnitude or consistently observed (56). Few MR studies have been conducted on the role of inflammation on cognition. Consistent with results presented here, our previous MR study examining the role of the same inflammatory markers (i.e., CRP, IL-6, IL-6R, sIL-6R, GlycA) on specific executive functions within the ALSPAC cohort (e.g., emotion recognition, working memory, response inhibition (56)) found little evidence of a potential causal effect. However, Pagoni et al. recently reported that other cytokines and chemokines (i.e., Eotaxin, IL-8, MCP1) may be causally related to lower fluid intelligence (and IL-4 with higher fluid intelligence) (57). The finding regarding sIL-6R and memory performance is novel and would align with convergent evidence that trans-signaling – in which sIL-6R plays a critical role – may be responsible for the deleterious effect of IL-6 on cognitive functioning (47, 108).

Interpreting the relationship between inflammation and cognitive task performance in population-based studies is difficult for several reasons. First, there is considerable heterogeneity in the type and breadth of cognitive abilities assessed across studies and there is a need for future studies to more uniformly include well-validated measures assessing individual differences [rather than detecting pathological states (e.g., dementia, epilepsy)] that characterize a broad range of cognitive functions (109). There is a similar need to measure and conceptualize the impact that inflammation has on other psychological functions that impact cognition (e.g., reward process, aversive process) – there is strong theoretical work and empirical data to support an indirect effect of inflammation on cognition via, for instance, dysregulated reward circuitry that impact performance on cognitive tasks via decreased motivation or increased fatigue (110). Moreover, there are a range of sociodemographic factors that may moderate the association between inflammation and cognition – prior work has found that inflammation and cognition may differ based on age and sex (111, 112). It is reasonable to assume, for instance, that modest increases in inflammation may exert a cumulative effect across the lifespan, and thus may only be detected later in life and/or in specific domains of cognition.

### 4.4 Anxiety

In the non-genetic analyses, circulating CRP levels was associated with a modestly increased likelihood of meeting criteria for anxiety disorders, although this association was substantially attenuated following adjustments for covariates. Prior research in population-based cohorts have found CRP to associated with an increased risk for anxiety disorders (113, 114), although results are inconsistent and other studies indicate that anxiety prospectively predicts an increase in circulating CRP levels (115). The MR analysis suggests a potential causal role of CRP on any anxiety disorder (which covers a broad range of anxiety-related conditions including panic disorder, social phobia, agoraphobia, GAD). Prior theory has primarily focused on anxiety as a cause of inflammation [see O’Donovan et al. for an excellent review (116)]; however, alternative theories suggest that inflammatory physiology is implicated in both sickness behaviors (e.g., anhedonia, social withdrawal) *and* anxiety arousal and alarm (117), which would align with the results presented here.

### 4.5 Limitations

In this study we used a large and broadly representative population-based sample, and we employed a triangulation of methods (non-genetic and genetic analyses) which increases confidence in the inferences drawn. Nevertheless, results should be considered in the context of the limitations of the study. First, although broadly representative, like other cohort studies (e.g., UK Biobank), the Lifelines cohort predominantly includes individuals of European descent and is less representative of individuals from low socioeconomic status (118), which consequentially limits the generalizability of findings. Second, analyses were not corrected for multiple comparisons. To check effects are not due to Type 1 errors, there is a need to replicate these findings in other cohorts. Moreover, as effect sizes reported are small and reflect associations in the general population, there is a need for studies to investigate whether there are sub-groups for whom these associations may be larger (e.g., older age, clinical populations). Third, in the genetic analysis the CRP GRS explained 1-4% of the variance in CRP [a level of variance consistent with similar analyses in the ALSPAC cohort (56)] and few cases of depression were observed in Lifelines [although the point prevalence of approximately 4% is consistent with reported population point prevalence estimates (119)]. It is possible that this limited our capacity to detect potential causal effects, were they small in magnitude or non-linear. Fourth, the CogState tasks used in the current study may not be optimal for detecting individual differences in healthy individuals, or even in some conditions such as depression; multiple studies have shown that the CogState tasks used in this study do not improve in successful antidepressant trials, even when improvement in other cognitive measures are observed (120–122). Fifth, although we include multiple instruments related to IL-6 (i.e., genetic variants related to IL-6 and sIL-6R levels), most instruments contain few genetic variants (≤ 3 SNPs) and genetic variants for IL-6 and sIL-6R overlap. While sIL-6R is involved in IL-6 trans-signaling, the overlap of SNPs makes it challenging to interpret the effect of these genetic variants on different immune phenotypes specifically (i.e., IL-6 levels vs IL-6 signaling). Future studies are needed to (1) better understand the biological role of these genetic variants and (2) develop instruments proxying specific IL-6 signaling pathways including IL-6 trans-signaling. Finally, it is worth considering that some instruments in the genetic analyses were associated with potential confounds, although it is unlikely that the small effects observed for some confounds substantially bias parameter estimates.

### 4.6 Conclusions

Genetic and non-genetic analyses provide consistent evidence for a modest effect of CRP on negative affect. Genetic analyses suggest that IL-6 signaling could be relevant for memory, and that the association between CRP and anxiety disorders could be causal. Overall, these results suggest that inflammation may affect a range of emotional states beyond the cardinal features of depression. However, given the small effect sizes and multiple tests conducted, future studies are required to investigate whether effects are moderated by sub-groups and whether these findings replicate in other cohorts.

## Data Availability

Conditions of data access can be found at www.lifelines-biobank.com/researchers/working-with-us.

## Acknowledgments

The Lifelines initiative has been made possible by subsidy from the Dutch Ministry of Health, Welfare and Sport, the Dutch Ministry of Economic Affairs, the University Medical Center Groningen (UMCG), Groningen University and the Provinces in the North of the Netherlands (Drenthe, Friesland, Groningen). The generation and management of GWAS genotype data for the Lifelines Cohort Study is supported by the UMCG Genetics Lifelines Initiative (UGLI). UGLI is partly supported by a Spinoza Grant from NWO, awarded to Cisca Wijmenga. The authors wish to acknowledge the services of the Lifelines Cohort Study, the contributing research centers delivering data to Lifelines, and all the study participants.

## Author Contributions

NMG, GMK, CH and CS conceptualized and designed the study. NMG conducted the non-genetic analysis. CS conducted the genetic analyses. NMG and CS wrote the first draft of the paper. All authors advised on the project/analysis and critically reviewed the final version of the manuscript. CH and GMK provided overall supervision for this project.

