## Supplementary Materials for "Role of Inflammation in Depressive and Anxiety Disorders, Affect, and Cognition: Genetic and Non-Genetic Findings in the Lifelines Cohort Study"

### Supplementary Material

### SUPPLEMENTARY LIFELINES GROUP AUTHOR INFORMATION

UMCG Genetics Lifelines Initiative (UGLI) group author: LifeLines Cohort Study

Raul Aguirre-Gamboa (1), Patrick Deelen (1), Lude Franke (1), Jan A Kuivenhoven (2), Esteban A Lopera Maya (1), Ilja M Nolte (3), Serena Sanna (1), Harold Snieder (3), Morris A Swertz (1), Peter M. Visscher (3,4), Judith M Vonk (3), Cisca Wijmenga (1), Naomi Wray (4)

(1) Department of Genetics, University of Groningen, University Medical Center Groningen, The Netherlands

(2) Department of Pediatrics, University of Groningen, University Medical Center Groningen, The Netherlands

(3) Department of Epidemiology, University of Groningen, University Medical Center Groningen, The Netherlands

(4) Institute for Molecular Bioscience, The University of Queensland, Brisbane, Queensland, Australia.

### SUPPLEMENTARY METHODS

##### *1.1. Cogstate Test Battery*

*Detection Task.* This is a reaction time task designed to assess psychomotor functioning and processing speed (Kuiper et al., 2017). In this task, participants attend to a card in the centre of the screen and respond to the question “Has the card turned face up” with “Yes” as soon as the card faces up. The task ends after 35 correct trials. The primary outcome is reaction time (ms) normalised using log10 transformation.

*Identification Task.* This is a reaction time task designed to measure visual attention (Kuiper et al., 2017). In this task, participants attend to a card in the centre of the screen and respond to the question “Is the card red?” with “Yes” or “No”. The task ends after 30 correct trials. The primary outcome is reaction time (ms) normalized using log10 transformation.

*One-back Task.* This task is designed to measure of attention and working memory (Kuiper et al., 2017). In this task, participants attend to a card in the centre of the screen and respond to the question “Is this card the same as that on the immediately previous trial?” with “Yes” or “No”. The task ends after 30 correct trials. The primary outcome is proportion of correct answers, normalized using arcsine transformation.

*One Card Learning Task.* This task is designed to measure visual learning and memory (Kuiper et al., 2017). In this task, participants attend to a card in the centre of the screen and respond to the question “Have you seen this card before in this task?” with “Yes” or “No”. The task ends after 42 trials. The primary outcome is proportion of correct answers normalized using arcsine transformation.

##### *1.2. Lifelines Genetics Data*

*CytoSNP*. In total, ~17,000 participants were genotyped in different batches using Illumina HumanCytoSNP-12v2.0 (n~16,500) and HumanCytoSNP-12v2.1 (n~500). Only probes present on both platforms were included. Genotyping was done using OptiCall and calls were refined using Beagle. CytoSNP originally used Genome Build 36, the probes were remapped to Genome Build 37 using SHRiMP2, with all probes are mapped on the forward strand. In total, 264,922 variants were present on both versions of the used HumanCytoSNP. Quality controls included excluding individuals with: (1) gender mismatches, (2) minimal or excessive heterozygosity, (3) duplicate sample identification, (4) missingness (call-rate < 95%), (5) non-Caucasian (determined by self-report in Lifelines phenotype database, Outlier [IBS] analysis, population stratification using Eigenstrat), (6) cryptic relationships (if a pair of samples were indicated as first-degree relatives using genetic similarity, the sample with the best genotyping quality was included). This resulted in 15,422 participants being available. Variants with minor allele frequency (MAF) of < 1%, call rate < 95%, or evidence for violations of Hardy-Weinberg Equilibrium ($p < 0.001$) were removed. Phasing was done using SHAPEIT2 and imputation using IMPUTE2 (combined reference panel of both genomes Genome of the Netherlands release 5 and 1000 Genomes phase1 v3 was used). For further details, please see: http://wiki-lifelines.web.rug.nl/doku.php?id=gwas.

*GSA [UGLI cohort].* In total, 38,030 participants were genotyped in 31 batches using the Infinium Global Screening Array® (GSA) MultiEthnic Disease version 1.0. In total, 691,072 variants were genotyped, of which 571,420 markers met quality control steps. Quality controls included excluding individuals with: (1) gender mismatches, (2) heterozygosity (>4 standard deviations from mean), (3) duplicate samples, (4) missingness (two-step process: first removed individuals > 20% missingness and then > 1% missingness). Participants were of Caucasian-ancestry, although PCA analysis detected 35 participants who were non-European. Variants which were monomorphic (MAF = 0), call rate < 1%, HWE ($p \leq 1 \times 10^{-6}$) were removed. A final set of 36,339 participants and 571,420 variants on autosomal and X chromosomes passed quality steps described above and were used for genetic imputation. Genetic imputation was done through the Sanger imputation service using the Haplotype Reference Consortium (http://www.haplotype-reference-consortium.org) panel. Following format instructions from the Sanger webpage (https://www.sanger.ac.uk/science/tools/sanger-imputation-service), 152 tri-allelic variants and 1608 insertions/deletions were removed. To facilitate researchers, Lifelines provide a list of non-European participants and a list of poorly imputed SNPs. For further details, please see: http://wiki-lifelines.web.rug.nl/lib/exe/fetch.php?media=qc_report_ugli_r1.pdf and http://wiki-lifelines.web.rug.nl/doku.php?id=ugli.

*Affymetrix [UGLI2 cohort].* In total, 29,166 participants were genotyped in 12 batches using the FinnGen Thermo Fisher Axiom® Custom array. Genotyping was done in Human Genome Build hg38. Quality controls included excluding individuals with: (1) sample mix-ups (using gender mismatches and pedigree concordance), (2) heterozygosity (> 4 SD from the mean), (3) duplicate sample, (4) missingness (two-step process: first removed individuals with > 20%

missingness and then > 3% missingness). A genetic relationship matrix was created for the 1000G cohort (without the admixed AMR population samples) (https://www.internationalgenome.org/) and used for principle-component-analysis (PCA) of up to 20 principal components to generate PC-loadings that were projected onto the UGLI2 cohort. The PC analysis of all 1000G superpopulations identified 142 non-Europeans (>4 SDs from centroid 1000G European population for first five PCs), and PC analysis of only 1000G European population identified 161 genetic outliers (>4 SDs from centroid of all UGLI2 samples for first two PCs). In this study, we removed non-European and genetic outliers from the dataset. Variants with MAF < 0.02%, call rate < 1%, HWE in all samples ($p<1x10^{-10}$), HWE in unrelated samples ($p<1x10^{-6}$; defined as no 1st or 2nd degree relations) were excluded. There were no SNPs with > 1% Mendelian errors across all parent-offspring pairs. Prior to imputation, genetic markers were lifted over to Genome Build GRCh37 and aligned with Haplotype Reference Consortium (HRC) v1.1 (http://www.haplotype-referenceconsortium.org/site). A final set of 28,250 samples and 462,731 markers on autosomal and X chromosomes passing quality check steps above were used for genetic imputation using the Sanger imputation service using HRC panel. For further details, please see: http://wiki-lifelines.web.rug.nl/lib/exe/fetch.php?media=qc_report_ugli2_release_1_-v1.pdf and http://wiki-lifelines.web.rug.nl/doku.php?id=ugli.

*Principal Components.* Lifelines provides genetic PCs for each chip separately, and PCs created using combined data (using all individuals). For the GRAMMAR method, PCs calculated on each chip separately were used. For unadjusted analysis and analysis removing related individuals, combined PCs were used.

##### *1.3. Details of GWAS and Instruments used to create GRS in Lifelines*

**Said et al. (2022). Meta analysis of two GWAS:**

> **CHARGE Consortium (Ligthart et al., 2018)** Circulating CRP was natural log transformed. Individuals were excluded from analyses if they had an auto-immune disease, were taking immune-modulating agents (if information was available) or had CRP ≥ 4 SD from the mean.
>
> **UK Biobank** Circulating CRP was natural log transformed, and individuals with extreme values (± 4 SD) from the mean were excluded. Individuals taking immune modulating drugs, who had auto-immune related conditions (1.8% sample), were removed.

**Ahluwalia et al. (2021)** Circulating IL-6 was natural log transformed. Only population-based samples or healthy controls from case-control studies were included.

**Sarwar et al. (2012)** Circulating IL-6 was natural log transformed. This instrument is composed of a single SNP (rs2228145) in the *IL6R* gene. The minor allele of this SNP (358A1a) is associated with increased sIL6R levels and decreased CRP levels[4,5]. Cell based experiments show that the minor allele decreases classical signalling [4]. Therefore, the effect of this SNP can be seen as a proxy for IL-6 activity. For more information on the biological function of this SNP, see [4,6].

**Borges et al. (2020)** Details not available.

**Rosa et al. (2019)** This SNP list was based on the Sun et al. (2018) sIL6R GWAS. The GWAS includes participants in good health. Individuals were excluded if they had a history of major disease (such as myocardial infarction, stroke, cancer, HIV, and hepatitis B or C) or and had a recent illness or infection. For more information, see[8]. Quality controls included exclusions for sex mismatches, low call rates, duplicate sample, extreme heterozygosity, and non-European descent.

##### *1.4. Adjustment for relatedness in genetic analyses*

To adjust for relatedness within each chip, two approaches were taken. First, we applied the GRAMMAR method *(*primary analysis)[9]: on each chip, we performed restricted estimated maximum likelihood predicting each outcome with the sparse genetic relationship matrix (GRM) and top 10 genetic principal components (PCs) as predictors. This was done using --*reml-pred-rand* in GCTA[10]. We extracted the residuals from each model and merged the data across chips (standardized GRS, residuals, age, sex). We then performed linear regressions to predict the outcome residuals using standardized GRS, age, sex, and chip (to adjust for potential differences between chips). Second, we re-ran analysis removing close relatives (secondary analysis) using *king-cutoff* [11] in Plink v2.0 [12,13] to identify relatives using conventional cut-offs for kinship coefficients (up to first-degree: 0.177; up to second-degree: 0.088; up to third-degree: 0.044).

##### *1.5. Creating Genetic Relationship Matrices (GRM)*

Using genotype (non-imputed) data, we created three GRM's (one for each chip). SNPs were used to create the GRM if they met the following criteria: MAF (0.01), call rate (0.95), HWE (1e-6), not multi-allelic. Independent SNPs were selected using --*indep-pairwise* [50 10 0.1] in Plink v1.9. Following this, a sparse GRM was created by replacing non-diagonal values <0.125 to 0 using --*make-bK* in GCTA[10]. A sparse matrix was created so that inclusion of the GRM will only adjust for recent relatedness.

#### SUPPLEMENTARY RESULTS

##### *2.1. GRS analysis*

Primary analyses were run adjusted for close relatedness within chips using a genetic relationship matrix (GRAMMAR method). Secondary analyses included (1) re-running the analysis unadjusted for relatedness within chips and (2) re-running the analysis removing close relatives within chips (up to first-degree, up to second-degree, up to third-degree). Conclusions are consistent across analyses.

##### *2.2. GRSs associations with exposures and potential confounders*

Linear regression models checked whether CRP GRS's predicted circulating levels of CRP in 23,607 Lifelines participants who had both genetic and CRP data available. The GRAMMAR method was used. All instruments had *F*-statistics >10 (158 for *cis* GRS, 1045 for genome-wide GRS), indicating adequate instrument strength[14,15]. *Cis* and genome-wide instruments explained 0.7% and 4.2% variance in CRP levels, respectively. The amount of variance explained by CRP GRS's in circulating CRP is consistent with our previous results in the ALSPAC cohort[16].

Linear regression models examined whether GRS's were associated with potential confounders. For primary instruments, there was evidence that the CRP *cis* instrument was associated with smoking ($p$=0.035) and weak evidence that the sIL6R instrument was associated with lower education attainment ($p$=0.061). For secondary instruments, there was evidence that the CRP genome-wide instrument was associated with BMI ($p$<0.001), smoking ($p$=0.0003) and lower education attainment ($p$=0.0004). Evidence for other GRS's was weak (Supplementary Table 3). Stronger evidence of associations between genome-wide instruments and potential confounders (consistent to what was reported in the previous ALSPAC study) highlights that *cis* variants may provide more valid instruments for MR [16].

#### SUPPLEMENTARY DISCUSSION

##### *3.1. Possible factors contributing to mixed MR results of CRP-depression*

Four factors which may contribute are CRP SNP selection, how depression was measured, statistical power, and selection bias. Regarding SNP selection, all previous studies (including the current study) used *cis* CRP SNPs; and previous studies included the same four *cis* SNPs (rs1205, rs1130864, rs3093077, rs3091244) as either a primary or secondary instrument. Thus, it is unlikely that SNP selection is driving discrepancy. Depression has been characterized differently across studies. This includes self-reported probable lifetime depression[17], hospitalization or death with depression[18], depression questionnaires/interviews assessed using PHQ-9 and HADS-D (coded continuously and categorically) [19,20] or MINI (current study), or a mixture of the above[21]. Different measures assess different symptoms of depression (e.g., the PHQ-9 includes somatic symptoms which are not included in measures such as HADS-D[20]). Therefore, if CRP differentially affects specific depression symptoms, this may contribute to mixed results observed. Selection bias should also be considered. For example, the study by Ye et al. (2021) which reported higher CRP to be associated with decreased risk of depression only included a subset of UK Biobank participants who responded to a mental health survey at follow-up, and consequently may be affected by selection bias.

#### SUPPLEMENTARY TABLES

***Table 1. Access to GWAS Data***

| GWAS Full Summary Statistics/Instruments | Access | URL *(if available online)* or author contact details |
|---|---|---|
| Said et al. (2022) | Requested from authors | https://www.ebi.ac.uk/gwas/studies/GCST90029070 |
| Ahluwalia et al. (2021) | Requested from authors | Corresponding authors:<br>Tarunveer Ahluwalia<br><br>Behrooz Alizadeh<br> |
| Sarwar et al. (2012) | Taken from OSF | osf.io/apme9/ |
| Borges et al. (2020) | IEU Open GWAS Project | gwas.mrcieu.ac.uk/datasets/met-d-GlycA/ |
| Rosa et al. (2019) | Available in paper supplementary | www.nature.com/articles/s41525-019-0097-4#Sec30 |

***Table 2. Details of GWAS used to create GRS***

| Phenotype | GWAS/ Instrument | Population | Cohort/ studies(*s*) | Covariates | Ages | N | Includes Lifelines (approximate % sample if applicable) | Ref |
|---|---|---|---|---|---|---|---|---|
| **CRP** | Said et al. (2022) | European ancestry | Meta-analysis of GWAS from CHARGE consortium (n=148,164) and UK Biobank (n=427,367). | CHARGE adjusted for age, sex, population structure, accounting for relatedness, if relevant. | CHARGE consortium cohorts range from *M* age of 9.9 to 86.6 years<br><br>UK Biobank age (M=56.5, SD=8.1) | 575,531 | Yes, Lifelines Cohort is in CHARGE GWAS; n ≤ **2.2%.** | 1 |
| **IL-6/IL-6R** | Ahluwalia et al. (2021) | European ancestry | 26 cohorts | Adjusted for age, sex, population substructure (through study-specific principal components) and/or study-specific site, when necessary. | Cohorts range from *M* age of 9.9 to 86.6 years | 52,654 | No | 2 |
| | Sarwar et al. (2012) Instrument | European ancestry (≥ 90%) | 16 studies | Unknown | Unknown | 27,185 | No | 3 |
| **GlycA** | Borges et al. (2020) | European | UK Biobank | Unknown | Unknown | 115,078 | No | N/A |
| **sIL6R** | Rosa et al. (2019) Instrument from Sun et al., (2018) GWAS on sIL6R. | European ancestry | INTERVAL study (UK) | Adjusted for sex, age, duration between blood draw and processing, first 3 ancestry principal components. | Cohorts *M* age is 44 years (*SD* = 14) | 3,301 | No | 7,8 |

***Table 3. Standardised GRS on potential confounders (GRAMMAR adjusted)***

| Standardised GRS | Outcome | Beta | p-value | LCI | UCI | N |
|---|---|---|---|---|---|---|
| CRP (said-cis) | BMI | -0.00399 | 0.6267 | -0.02005 | 0.01208 | 58680 |
| CRP (said-cis) | Current smoke status | 0.00272 | 0.0347 | 0.00020 | 0.00524 | 57935 |
| CRP (said-cis) | Education attainment | 0.00163 | 0.3885 | -0.00207 | 0.00533 | 58094 |
| GlycA (borges) | BMI | 0.00541 | 0.5094 | -0.01066 | 0.02147 | 58680 |
| GlycA (borges) | Current smoke status | 0.00140 | 0.2777 | -0.00113 | 0.00392 | 57935 |
| GlycA (borges) | Education attainment | 0.00068 | 0.7169 | -0.00302 | 0.00439 | 58094 |
| IL-6R (ahluwalia-cis) | BMI | -0.00305 | 0.7102 | -0.01911 | 0.01302 | 58680 |
| IL-6R (ahluwalia-cis) | Current smoke status | 0.00156 | 0.2271 | -0.00097 | 0.00408 | 57935 |
| IL-6R (ahluwalia-cis) | Education attainment | 0.00199 | 0.2928 | -0.00172 | 0.00569 | 58094 |
| sIL6R (rosa) | BMI | 0.00669 | 0.4141 | -0.00937 | 0.02276 | 58680 |
| sIL6R (rosa) | Current smoke status | 0.00159 | 0.2173 | -0.00094 | 0.00411 | 57935 |
| sIL6R (rosa) | Education attainment | 0.00354 | 0.0608 | -0.00016 | 0.00724 | 58094 |
| IL-6R (sarwar) | BMI | 0.00172 | 0.8339 | -0.01435 | 0.01779 | 58680 |
| IL-6R (sarwar) | Current smoke status | 0.00124 | 0.3371 | -0.00129 | 0.00376 | 57935 |
| IL-6R (sarwar) | Education attainment | 0.00192 | 0.3103 | -0.00179 | 0.00562 | 58094 |
| CRP (said-genome-wide) | BMI | 0.04719 | <0.0001 | 0.03113 | 0.06324 | 58680 |
| CRP (said-genome-wide) | Current smoke status | 0.00465 | 0.0003 | 0.00213 | 0.00717 | 57935 |
| CRP (said-genome-wide) | Education attainment | 0.00671 | 0.0004 | 0.00301 | 0.01041 | 58094 |
| IL-6 (ahluwalia-genome-wide) | BMI | -0.00408 | 0.6194 | -0.02016 | 0.01201 | 58680 |
| IL-6 (ahluwalia-genome-wide) | Current smoke status | 0.00125 | 0.3325 | -0.00128 | 0.00378 | 57935 |
| IL-6 (ahluwalia-genome-wide) | Education attainment | 0.00211 | 0.2655 | -0.00160 | 0.00581 | 58094 |

***Table 4. Standardised GRS on standardised outcomes (GRAMMAR adjusted).***

| Standardised GRS | Outcome | Beta | p-value | LCI | UCI | N |
|---|---|---|---|---|---|---|
| CRP (said-cis) | Identify Task – reaction time | -0.00251 | 0.502 | -0.00986 | 0.00483 | 35729 |
| CRP (said-cis) | Detect Task – reaction time | 0.00389 | 0.355 | -0.00435 | 0.01213 | 35300 |
| CRP (said-cis) | OCL Task - accuracy | -0.00566 | 0.153 | -0.01341 | 0.00210 | 36783 |
| CRP (said-cis) | One-back Task - accuracy | -0.00646 | 0.159 | -0.01547 | 0.00254 | 36349 |
| CRP (said-cis) | RFFT | 0.00460 | 0.217 | -0.00271 | 0.01191 | 36563 |
| CRP (said-cis) | PANAS – negative | 0.00646 | 0.035 | 0.00046 | 0.01247 | 57946 |
| CRP (said-cis) | PANAS - positive | 0.00405 | 0.192 | -0.00203 | 0.01014 | 57946 |
| CRP (said-cis) | Mini depression | 0.00079 | 0.217 | -0.00046 | 0.00204 | 56860 |
| CRP (said-cis) | Mini anxiety | 0.00198 | 0.037 | 0.00012 | 0.00384 | 57047 |
| CRP (said-cis) | Mini MDD | 0.00037 | 0.486 | -0.00067 | 0.00140 | 57047 |
| CRP (said-cis) | Mini GAD | 0.00101 | 0.164 | -0.00041 | 0.00243 | 57047 |
| GlycA (borges) | Identify Task – reaction time | -0.00002 | 0.996 | -0.00737 | 0.00733 | 35729 |
| GlycA (borges) | Detect Task – reaction time | 0.00093 | 0.826 | -0.00732 | 0.00918 | 35300 |
| GlycA (borges) | OCL Task - accuracy | 0.00232 | 0.558 | -0.00544 | 0.01007 | 36783 |
| GlycA (borges) | One-back Task - accuracy | 0.00110 | 0.810 | -0.00791 | 0.01011 | 36349 |
| GlycA (borges) | RFFT | -0.00541 | 0.148 | -0.01273 | 0.00191 | 36563 |
| GlycA (borges) | PANAS – negative | 0.00603 | 0.049 | 0.00002 | 0.01203 | 57946 |
| GlycA (borges) | PANAS - positive | -0.00185 | 0.552 | -0.00794 | 0.00424 | 57946 |
| GlycA (borges) | Mini depression | 0.00087 | 0.172 | -0.00038 | 0.00212 | 56860 |
| GlycA (borges) | Mini anxiety | 0.00021 | 0.821 | -0.00164 | 0.00207 | 57047 |
| GlycA (borges) | Mini MDD | 0.00110 | 0.036 | 0.00007 | 0.00213 | 57047 |
| GlycA (borges) | Mini GAD | 0.00053 | 0.465 | -0.00089 | 0.00195 | 57047 |
| IL-6R (ahluwalia-cis) | Identify Task – reaction time | -0.00504 | 0.179 | -0.01239 | 0.00232 | 35729 |
| IL-6R (ahluwalia-cis) | Detect Task – reaction time | -0.00144 | 0.733 | -0.00970 | 0.00682 | 35300 |
| IL-6R (ahluwalia-cis) | OCL Task - accuracy | -0.00394 | 0.320 | -0.01170 | 0.00383 | 36783 |
| IL-6R (ahluwalia-cis) | One-back Task - accuracy | 0.00171 | 0.710 | -0.00730 | 0.01072 | 36349 |
| IL-6R (ahluwalia-cis) | RFFT | -0.00395 | 0.291 | -0.01127 | 0.00338 | 36563 |
| IL-6R (ahluwalia-cis) | PANAS – negative | 0.00348 | 0.257 | -0.00253 | 0.00949 | 57946 |
| IL-6R (ahluwalia-cis) | PANAS - positive | -0.00099 | 0.750 | -0.00708 | 0.00510 | 57946 |
| IL-6R (ahluwalia-cis) | Mini depression | -0.00024 | 0.708 | -0.00149 | 0.00101 | 56860 |

| | | | | | | |
|---|---|---|---|---|---|---|
| IL-6R (ahluwalia-cis) | Mini anxiety | -0.00022 | 0.819 | -0.00208 | 0.00164 | 57047 |
| IL-6R (ahluwalia-cis) | Mini MDD | -0.00002 | 0.964 | -0.00106 | 0.00101 | 57047 |
| IL-6R (ahluwalia-cis) | Mini GAD | 0.00013 | 0.857 | -0.00129 | 0.00155 | 57047 |
| sIL6R (rosa) | Identify Task – reaction time | -0.00423 | 0.259 | -0.01158 | 0.00312 | 35729 |
| sIL6R (rosa) | Detect Task – reaction time | -0.00080 | 0.849 | -0.00905 | 0.00746 | 35300 |
| sIL6R (rosa) | OCL Task - accuracy | -0.00936 | 0.018 | -0.01712 | -0.00161 | 36783 |
| sIL6R (rosa) | One-back Task - accuracy | -0.00442 | 0.336 | -0.01343 | 0.00459 | 36349 |
| sIL6R (rosa) | RFFT | -0.00063 | 0.867 | -0.00793 | 0.00668 | 36563 |
| sIL6R (rosa) | PANAS – negative | 0.00459 | 0.134 | -0.00142 | 0.01060 | 57946 |
| sIL6R (rosa) | PANAS - positive | -0.00228 | 0.462 | -0.00837 | 0.00381 | 57946 |
| sIL6R (rosa) | Mini depression | -0.00057 | 0.369 | -0.00182 | 0.00068 | 56860 |
| sIL6R (rosa) | Mini anxiety | -0.00005 | 0.960 | -0.00191 | 0.00181 | 57047 |
| sIL6R (rosa) | Mini MDD | -0.00009 | 0.863 | -0.00112 | 0.00094 | 57047 |
| sIL6R (rosa) | Mini GAD | 0.00009 | 0.901 | -0.00133 | 0.00151 | 57047 |
| IL-6R (sarwar) | Identify Task – reaction time | -0.00434 | 0.246 | -0.01168 | 0.00300 | 35729 |
| IL-6R (sarwar) | Detect Task – reaction time | -0.00183 | 0.663 | -0.01007 | 0.00641 | 35300 |
| IL-6R (sarwar) | OCL Task - accuracy | -0.00588 | 0.137 | -0.01363 | 0.00187 | 36783 |
| IL-6R (sarwar) | One-back Task - accuracy | 0.00117 | 0.798 | -0.00782 | 0.01017 | 36349 |
| IL-6R (sarwar) | RFFT | -0.00047 | 0.899 | -0.00779 | 0.00684 | 36563 |
| IL-6R (sarwar) | PANAS – negative | 0.00373 | 0.223 | -0.00227 | 0.00974 | 57946 |
| IL-6R (sarwar) | PANAS - positive | -0.00443 | 0.154 | -0.01052 | 0.00166 | 57946 |
| IL-6R (sarwar) | Mini depression | -0.00007 | 0.915 | -0.00132 | 0.00118 | 56860 |
| IL-6R (sarwar) | Mini anxiety | 0.00029 | 0.762 | -0.00157 | 0.00215 | 57047 |
| IL-6R (sarwar) | Mini MDD | 0.00003 | 0.948 | -0.00100 | 0.00107 | 57047 |
| IL-6R (sarwar) | Mini GAD | 0.00034 | 0.637 | -0.00108 | 0.00176 | 57047 |
| CRP (said-genome-wide) | Identify Task – reaction time | -0.00421 | 0.260 | -0.01155 | 0.00312 | 35729 |
| CRP (said-genome-wide) | Detect Task – reaction time | 0.00185 | 0.659 | -0.00637 | 0.01007 | 35300 |
| CRP (said-genome-wide) | OCL Task - accuracy | 0.00360 | 0.362 | -0.00414 | 0.01133 | 36783 |
| CRP (said-genome-wide) | One-back Task - accuracy | 0.00429 | 0.349 | -0.00468 | 0.01327 | 36349 |
| CRP (said-genome-wide) | RFFT | 0.00290 | 0.437 | -0.00442 | 0.01022 | 36563 |
| CRP (said-genome-wide) | PANAS – negative | -0.00221 | 0.471 | -0.00821 | 0.00380 | 57946 |
| CRP (said-genome-wide) | PANAS - positive | -0.00058 | 0.852 | -0.00666 | 0.00551 | 57946 |

| | | | | | | |
|---|---|---|---|---|---|---|
| CRP (said-genome-wide) | Mini depression | 0.00060 | 0.350 | -0.00065 | 0.00184 | 56860 |
| CRP (said-genome-wide) | Mini anxiety | 0.00215 | 0.023 | 0.00029 | 0.00401 | 57047 |
| CRP (said-genome-wide) | Mini MDD | 0.00031 | 0.561 | -0.00072 | 0.00134 | 57047 |
| CRP (said-genome-wide) | Mini GAD | 0.00078 | 0.279 | -0.00063 | 0.00220 | 57047 |
| IL-6 (ahluwalia-genome-wide) | Identify Task – reaction time | -0.00463 | 0.217 | -0.01199 | 0.00272 | 35729 |
| IL-6 (ahluwalia-genome-wide) | Detect Task – reaction time | -0.00143 | 0.735 | -0.00969 | 0.00684 | 35300 |
| IL-6 (ahluwalia-genome-wide) | OCL Task - accuracy | -0.00390 | 0.325 | -0.01166 | 0.00387 | 36783 |
| IL-6 (ahluwalia-genome-wide) | One-back Task - accuracy | 0.00157 | 0.733 | -0.00744 | 0.01059 | 36349 |
| IL-6 (ahluwalia-genome-wide) | RFFT | -0.00451 | 0.228 | -0.01184 | 0.00283 | 36563 |
| IL-6 (ahluwalia-genome-wide) | PANAS – negative | 0.00432 | 0.159 | -0.00169 | 0.01033 | 57946 |
| IL-6 (ahluwalia-genome-wide) | PANAS - positive | 0.00009 | 0.976 | -0.00600 | 0.00619 | 57946 |
| IL-6 (ahluwalia-genome-wide) | Mini depression | -0.00011 | 0.865 | -0.00136 | 0.00114 | 56860 |
| IL-6 (ahluwalia-genome-wide) | Mini anxiety | 0.00015 | 0.875 | -0.00171 | 0.00201 | 57047 |
| IL-6 (ahluwalia-genome-wide) | Mini MDD | 0.00002 | 0.969 | -0.00101 | 0.00105 | 57047 |
| IL-6 (ahluwalia-genome-wide) | Mini GAD | 0.00029 | 0.693 | -0.00113 | 0.00171 | 57047 |

***Table 5. Standardised GRS on binary outcomes (unadjusted analysis)***

| Standardised GRS | Outcome | OR | p-value | LCI | UCI | N |
|---|---|---|---|---|---|---|
| CRP (said-cis) | Mini depression | 1.034 | 0.185 | 0.984 | 1.085 | 56860 |
| CRP (said-cis) | Mini anxiety | 1.035 | 0.034 | 1.003 | 1.068 | 57047 |
| CRP (said-cis) | Mini MDD | 1.022 | 0.487 | 0.961 | 1.087 | 57047 |
| CRP (said-cis) | Mini GAD | 1.030 | 0.175 | 0.987 | 1.075 | 57047 |
| GlycA (borges) | Mini depression | 1.031 | 0.224 | 0.982 | 1.083 | 56860 |
| GlycA (borges) | Mini anxiety | 1.003 | 0.876 | 0.971 | 1.035 | 57047 |
| GlycA (borges) | Mini MDD | 1.068 | 0.037 | 1.004 | 1.137 | 57047 |
| GlycA (borges) | Mini GAD | 1.016 | 0.481 | 0.973 | 1.060 | 57047 |
| IL-6R (ahluwalia) | Mini depression | 0.990 | 0.693 | 0.942 | 1.040 | 56860 |
| IL-6R (ahluwalia) | Mini anxiety | 0.994 | 0.731 | 0.963 | 1.027 | 57047 |
| IL-6R (ahluwalia) | Mini MDD | 0.999 | 0.979 | 0.939 | 1.063 | 57047 |
| IL-6R (ahluwalia) | Mini GAD | 1.001 | 0.972 | 0.958 | 1.045 | 57047 |
| sIL6R (rosa) | Mini depression | 0.976 | 0.325 | 0.929 | 1.025 | 56860 |
| sIL6R (rosa) | Mini anxiety | 0.997 | 0.854 | 0.966 | 1.029 | 57047 |
| sIL6R (rosa) | Mini MDD | 0.995 | 0.871 | 0.935 | 1.059 | 57047 |
| sIL6R (rosa) | Mini GAD | 0.999 | 0.963 | 0.957 | 1.043 | 57047 |
| IL-6R (sarwar) | Mini depression | 0.996 | 0.887 | 0.948 | 1.047 | 56860 |
| IL-6R (sarwar) | Mini anxiety | 1.003 | 0.861 | 0.971 | 1.035 | 57047 |
| IL-6R (sarwar) | Mini MDD | 1.003 | 0.930 | 0.942 | 1.067 | 57047 |
| IL-6R (sarwar) | Mini GAD | 1.007 | 0.752 | 0.964 | 1.051 | 57047 |
| CRP (said-genome-wide) | Mini depression | 1.024 | 0.346 | 0.975 | 1.076 | 56860 |
| CRP (said-genome-wide) | Mini anxiety | 1.040 | 0.016 | 1.007 | 1.074 | 57047 |
| CRP (said-genome-wide) | Mini MDD | 1.016 | 0.627 | 0.954 | 1.081 | 57047 |
| CRP (said-genome-wide) | Mini GAD | 1.025 | 0.254 | 0.982 | 1.071 | 57047 |
| IL-6 (ahluwalia-genome-wide) | Mini depression | 0.995 | 0.832 | 0.947 | 1.045 | 56860 |
| IL-6 (ahluwalia-genome-wide) | Mini anxiety | 0.998 | 0.892 | 0.966 | 1.030 | 57047 |
| IL-6 (ahluwalia-genome-wide) | Mini MDD | 1.004 | 0.888 | 0.944 | 1.069 | 57047 |
| IL-6 (ahluwalia-genome-wide) | Mini GAD | 1.003 | 0.897 | 0.960 | 1.047 | 57047 |

***Table 6. Standardised GRS on continuous outcomes (unadjusted analysis)***

| Standardised GRS | Standardised Outcomes | Beta | p-value | LCI | UCI | N |
|---|---|---|---|---|---|---|
| CRP (said-cis) | Identify Task – reaction time | -0.00321 | 0.492 | -0.01235 | 0.00594 | 35729 |
| CRP (said-cis) | Detect Task – reaction time | 0.00437 | 0.359 | -0.00497 | 0.01372 | 35300 |
| CRP (said-cis) | OCL Task - accuracy | -0.00740 | 0.146 | -0.01737 | 0.00257 | 36783 |
| CRP (said-cis) | One-back Task - accuracy | -0.00627 | 0.219 | -0.01626 | 0.00372 | 36349 |
| CRP (said-cis) | RFFT | 0.00514 | 0.299 | -0.00455 | 0.01483 | 36563 |
| CRP (said-cis) | PANAS – negative | 0.00805 | 0.046 | 0.00016 | 0.01594 | 57946 |
| CRP (said-cis) | PANAS - positive | 0.00535 | 0.191 | -0.00266 | 0.01337 | 57946 |
| GlycA (borges) | Identify Task – reaction time | -0.00069 | 0.882 | -0.00984 | 0.00846 | 35729 |
| GlycA (borges) | Detect Task – reaction time | 0.00189 | 0.692 | -0.00747 | 0.01125 | 35300 |
| GlycA (borges) | OCL Task - accuracy | 0.00307 | 0.547 | -0.00691 | 0.01304 | 36783 |
| GlycA (borges) | One-back Task - accuracy | 0.00083 | 0.870 | -0.00916 | 0.01083 | 36349 |
| GlycA (borges) | RFFT | -0.00652 | 0.188 | -0.01623 | 0.00319 | 36563 |
| GlycA (borges) | PANAS – negative | 0.00916 | 0.023 | 0.00126 | 0.01706 | 57946 |
| GlycA (borges) | PANAS - positive | -0.00307 | 0.453 | -0.01109 | 0.00495 | 57946 |
| IL-6R (ahluwalia-cis) | Identify Task – reaction time | -0.00574 | 0.219 | -0.01489 | 0.00341 | 35729 |
| IL-6R (ahluwalia-cis) | Detect Task – reaction time | -0.00084 | 0.860 | -0.01021 | 0.00853 | 35300 |
| IL-6R (ahluwalia-cis) | OCL Task - accuracy | -0.00504 | 0.323 | -0.01502 | 0.00495 | 36783 |
| IL-6R (ahluwalia-cis) | One-back Task - accuracy | 0.00143 | 0.780 | -0.00857 | 0.01142 | 36349 |
| IL-6R (ahluwalia-cis) | RFFT | -0.00609 | 0.219 | -0.01580 | 0.00362 | 36563 |
| IL-6R (ahluwalia-cis) | PANAS – negative | 0.00402 | 0.318 | -0.00387 | 0.01192 | 57946 |
| IL-6R (ahluwalia-cis) | PANAS - positive | -0.00156 | 0.704 | -0.00957 | 0.00646 | 57946 |
| sIL6R (rosa) | Identify Task – reaction time | -0.00482 | 0.301 | -0.01397 | 0.00432 | 35729 |
| sIL6R (rosa) | Detect Task – reaction time | 0.00002 | 0.996 | -0.00933 | 0.00938 | 35300 |
| sIL6R (rosa) | OCL Task - accuracy | -0.01197 | 0.019 | -0.02195 | -0.00200 | 36783 |
| sIL6R (rosa) | One-back Task - accuracy | -0.00545 | 0.285 | -0.01545 | 0.00454 | 36349 |
| sIL6R (rosa) | RFFT | -0.00128 | 0.795 | -0.01097 | 0.00840 | 36563 |
| sIL6R (rosa) | PANAS – negative | 0.00567 | 0.159 | -0.00223 | 0.01357 | 57946 |
| sIL6R (rosa) | PANAS - positive | -0.00293 | 0.474 | -0.01095 | 0.00509 | 57946 |
| IL-6R (sarwar) | Identify Task – reaction time | -0.00497 | 0.286 | -0.01410 | 0.00417 | 35729 |
| IL-6R (sarwar) | Detect Task – reaction time | -0.00120 | 0.802 | -0.01054 | 0.00815 | 35300 |

| | | | | | | |
|---|---|---|---|---|---|---|
| IL-6R (sarwar) | OCL Task - accuracy | -0.00772 | 0.129 | -0.01769 | 0.00224 | 36783 |
| IL-6R (sarwar) | One-back Task - accuracy | 0.00065 | 0.899 | -0.00933 | 0.01062 | 36349 |
| IL-6R (sarwar) | RFFT | -0.00186 | 0.706 | -0.01156 | 0.00783 | 36563 |
| IL-6R (sarwar) | PANAS – negative | 0.00441 | 0.274 | -0.00349 | 0.01230 | 57946 |
| IL-6R (sarwar) | PANAS - positive | -0.00602 | 0.141 | -0.01404 | 0.00200 | 57946 |
| CRP (said-genome-wide) | Identify Task – reaction time | -0.00477 | 0.305 | -0.01389 | 0.00435 | 35729 |
| CRP (said-genome-wide) | Detect Task – reaction time | 0.00253 | 0.595 | -0.00679 | 0.01185 | 35300 |
| CRP (said-genome-wide) | OCL Task - accuracy | 0.00518 | 0.307 | -0.00477 | 0.01512 | 36783 |
| CRP (said-genome-wide) | One-back Task - accuracy | 0.00607 | 0.232 | -0.00388 | 0.01603 | 36349 |
| CRP (said-genome-wide) | RFFT | 0.00498 | 0.314 | -0.00471 | 0.01468 | 36563 |
| CRP (said-genome-wide) | PANAS – negative | -0.00411 | 0.308 | -0.01199 | 0.00378 | 57946 |
| CRP (said-genome-wide) | PANAS - positive | -0.00006 | 0.989 | -0.00807 | 0.00795 | 57946 |
| IL-6 (ahluwalia-genome-wide) | Identify Task – reaction time | -0.00580 | 0.215 | -0.01498 | 0.00338 | 35729 |
| IL-6 (ahluwalia-genome-wide) | Detect Task – reaction time | -0.00149 | 0.756 | -0.01089 | 0.00791 | 35300 |
| IL-6 (ahluwalia-genome-wide) | OCL Task - accuracy | -0.00400 | 0.434 | -0.01401 | 0.00602 | 36783 |
| IL-6 (ahluwalia-genome-wide) | One-back Task - accuracy | 0.00133 | 0.794 | -0.00870 | 0.01136 | 36349 |
| IL-6 (ahluwalia-genome-wide) | RFFT | -0.00604 | 0.225 | -0.01578 | 0.00371 | 36563 |
| IL-6 (ahluwalia-genome-wide) | PANAS – negative | 0.00423 | 0.295 | -0.00369 | 0.01216 | 57946 |
| IL-6 (ahluwalia-genome-wide) | PANAS - positive | 0.00041 | 0.921 | -0.00764 | 0.00846 | 57946 |

***Table 7. Standardised GRS on binary outcomes (1st degree relatives removed)***

| Standardised GRS | Outcome | OR | p-value | LCI | UCI | N |
|---|---|---|---|---|---|---|
| CRP (said-cis) | Mini depression | 1.048 | 0.084 | 0.994 | 1.104 | 49183 |
| CRP (said-cis) | Mini anxiety | 1.035 | 0.048 | 1.000 | 1.071 | 49348 |
| CRP (said-cis) | Mini MDD | 1.041 | 0.238 | 0.974 | 1.112 | 49348 |
| CRP (said-cis) | Mini GAD | 1.036 | 0.137 | 0.989 | 1.085 | 49348 |
| GlycA (borges) | Mini depression | 1.046 | 0.096 | 0.992 | 1.103 | 49183 |
| GlycA (borges) | Mini anxiety | 1.016 | 0.364 | 0.982 | 1.051 | 49348 |
| GlycA (borges) | Mini MDD | 1.083 | 0.019 | 1.013 | 1.158 | 49348 |
| GlycA (borges) | Mini GAD | 1.046 | 0.056 | 0.999 | 1.096 | 49348 |
| IL-6R (ahluwalia) | Mini depression | 1.005 | 0.858 | 0.953 | 1.059 | 49183 |
| IL-6R (ahluwalia) | Mini anxiety | 1.001 | 0.973 | 0.967 | 1.035 | 49348 |
| IL-6R (ahluwalia) | Mini MDD | 1.001 | 0.974 | 0.936 | 1.070 | 49348 |
| IL-6R (ahluwalia) | Mini GAD | 1.006 | 0.816 | 0.960 | 1.053 | 49348 |
| sIL6R (rosa) | Mini depression | 0.987 | 0.639 | 0.936 | 1.041 | 49183 |
| sIL6R (rosa) | Mini anxiety | 0.998 | 0.921 | 0.965 | 1.033 | 49348 |
| sIL6R (rosa) | Mini MDD | 0.997 | 0.929 | 0.932 | 1.066 | 49348 |
| sIL6R (rosa) | Mini GAD | 1.003 | 0.903 | 0.957 | 1.050 | 49348 |
| IL-6R (sarwar) | Mini depression | 1.008 | 0.769 | 0.956 | 1.063 | 49183 |
| IL-6R (sarwar) | Mini anxiety | 1.011 | 0.534 | 0.977 | 1.046 | 49348 |
| IL-6R (sarwar) | Mini MDD | 1.005 | 0.880 | 0.940 | 1.074 | 49348 |
| IL-6R (sarwar) | Mini GAD | 1.013 | 0.594 | 0.967 | 1.061 | 49348 |
| CRP (said-genome-wide) | Mini depression | 1.038 | 0.172 | 0.984 | 1.095 | 49183 |
| CRP (said-genome-wide) | Mini anxiety | 1.038 | 0.032 | 1.003 | 1.074 | 49348 |
| CRP (said-genome-wide) | Mini MDD | 1.027 | 0.429 | 0.961 | 1.098 | 49348 |
| CRP (said-genome-wide) | Mini GAD | 1.028 | 0.246 | 0.981 | 1.077 | 49348 |
| IL-6 (ahluwalia-genome-wide) | Mini depression | 1.010 | 0.701 | 0.958 | 1.066 | 49183 |
| IL-6 (ahluwalia-genome-wide) | Mini anxiety | 1.004 | 0.802 | 0.970 | 1.039 | 49348 |
| IL-6 (ahluwalia-genome-wide) | Mini MDD | 1.008 | 0.820 | 0.942 | 1.077 | 49348 |
| IL-6 (ahluwalia-genome-wide) | Mini GAD | 1.008 | 0.733 | 0.962 | 1.056 | 49348 |

***Table 8. Standardised GRS on binary outcomes (2nd degree relatives removed)***

| Standardised GRS | Outcome | OR | p-value | LCI | UCI | N |
|---|---|---|---|---|---|---|
| CRP (said-cis) | Mini depression | 1.051 | 0.064 | 0.997 | 1.109 | 48493 |
| CRP (said-cis) | Mini anxiety | 1.037 | 0.039 | 1.002 | 1.073 | 48656 |
| CRP (said-cis) | Mini MDD | 1.045 | 0.195 | 0.977 | 1.118 | 48656 |
| CRP (said-cis) | Mini GAD | 1.036 | 0.140 | 0.988 | 1.085 | 48656 |
| GlycA (borges) | Mini depression | 1.046 | 0.100 | 0.991 | 1.103 | 48493 |
| GlycA (borges) | Mini anxiety | 1.016 | 0.355 | 0.982 | 1.052 | 48656 |
| GlycA (borges) | Mini MDD | 1.085 | 0.018 | 1.014 | 1.160 | 48656 |
| GlycA (borges) | Mini GAD | 1.047 | 0.056 | 0.999 | 1.097 | 48656 |
| IL-6R (ahluwalia) | Mini depression | 1.001 | 0.985 | 0.948 | 1.055 | 48493 |
| IL-6R (ahluwalia) | Mini anxiety | 1.002 | 0.921 | 0.968 | 1.037 | 48656 |
| IL-6R (ahluwalia) | Mini MDD | 0.989 | 0.751 | 0.924 | 1.058 | 48656 |
| IL-6R (ahluwalia) | Mini GAD | 1.000 | 0.999 | 0.954 | 1.048 | 48656 |
| sIL6R (rosa) | Mini depression | 0.985 | 0.591 | 0.934 | 1.040 | 48493 |
| sIL6R (rosa) | Mini anxiety | 1.000 | 0.993 | 0.966 | 1.035 | 48656 |
| sIL6R (rosa) | Mini MDD | 0.991 | 0.790 | 0.926 | 1.060 | 48656 |
| sIL6R (rosa) | Mini GAD | 1.000 | 0.990 | 0.955 | 1.048 | 48656 |
| IL-6R (sarwar) | Mini depression | 1.004 | 0.885 | 0.952 | 1.059 | 48493 |
| IL-6R (sarwar) | Mini anxiety | 1.012 | 0.492 | 0.978 | 1.047 | 48656 |
| IL-6R (sarwar) | Mini MDD | 0.995 | 0.879 | 0.930 | 1.064 | 48656 |
| IL-6R (sarwar) | Mini GAD | 1.007 | 0.776 | 0.961 | 1.055 | 48656 |
| CRP (said-genome-wide) | Mini depression | 1.044 | 0.117 | 0.989 | 1.101 | 48493 |
| CRP (said-genome-wide) | Mini anxiety | 1.040 | 0.026 | 1.005 | 1.077 | 48656 |
| CRP (said-genome-wide) | Mini MDD | 1.032 | 0.357 | 0.965 | 1.105 | 48656 |
| CRP (said-genome-wide) | Mini GAD | 1.029 | 0.237 | 0.982 | 1.078 | 48656 |
| IL-6 (ahluwalia-genome-wide) | Mini depression | 1.006 | 0.836 | 0.953 | 1.061 | 48493 |
| IL-6 (ahluwalia-genome-wide) | Mini anxiety | 1.006 | 0.737 | 0.972 | 1.041 | 48656 |
| IL-6 (ahluwalia-genome-wide) | Mini MDD | 0.995 | 0.889 | 0.930 | 1.065 | 48656 |
| IL-6 (ahluwalia-genome-wide) | Mini GAD | 1.003 | 0.900 | 0.957 | 1.051 | 48656 |

***Table 9. Standardised GRS on binary outcomes (3rd degree relatives removed)***

| Standardised GRS | Outcome | OR | p-value | LCI | UCI | N |
|---|---|---|---|---|---|---|
| CRP (said-cis) | Mini depression | 1.047 | 0.094 | 0.992 | 1.105 | 47136 |
| CRP (said-cis) | Mini anxiety | 1.035 | 0.052 | 1.000 | 1.072 | 47295 |
| CRP (said-cis) | Mini MDD | 1.039 | 0.265 | 0.971 | 1.112 | 47295 |
| CRP (said-cis) | Mini GAD | 1.031 | 0.210 | 0.983 | 1.081 | 47295 |
| GlycA (borges) | Mini depression | 1.037 | 0.190 | 0.982 | 1.095 | 47136 |
| GlycA (borges) | Mini anxiety | 1.018 | 0.321 | 0.983 | 1.054 | 47295 |
| GlycA (borges) | Mini MDD | 1.066 | 0.067 | 0.996 | 1.141 | 47295 |
| GlycA (borges) | Mini GAD | 1.046 | 0.064 | 0.997 | 1.097 | 47295 |
| IL-6R (ahluwalia) | Mini depression | 1.001 | 0.975 | 0.948 | 1.057 | 47136 |
| IL-6R (ahluwalia) | Mini anxiety | 1.003 | 0.858 | 0.969 | 1.039 | 47295 |
| IL-6R (ahluwalia) | Mini MDD | 0.986 | 0.682 | 0.920 | 1.055 | 47295 |
| IL-6R (ahluwalia) | Mini GAD | 1.011 | 0.657 | 0.964 | 1.060 | 47295 |
| sIL6R (rosa) | Mini depression | 0.985 | 0.590 | 0.933 | 1.040 | 47136 |
| sIL6R (rosa) | Mini anxiety | 0.999 | 0.937 | 0.964 | 1.034 | 47295 |
| sIL6R (rosa) | Mini MDD | 0.983 | 0.615 | 0.917 | 1.052 | 47295 |
| sIL6R (rosa) | Mini GAD | 1.006 | 0.808 | 0.959 | 1.055 | 47295 |
| IL-6R (sarwar) | Mini depression | 1.001 | 0.965 | 0.948 | 1.057 | 47136 |
| IL-6R (sarwar) | Mini anxiety | 1.010 | 0.562 | 0.976 | 1.046 | 47295 |
| IL-6R (sarwar) | Mini MDD | 0.988 | 0.727 | 0.922 | 1.058 | 47295 |
| IL-6R (sarwar) | Mini GAD | 1.014 | 0.579 | 0.966 | 1.063 | 47295 |
| CRP (said-genome-wide) | Mini depression | 1.039 | 0.171 | 0.984 | 1.097 | 47136 |
| CRP (said-genome-wide) | Mini anxiety | 1.040 | 0.027 | 1.004 | 1.078 | 47295 |
| CRP (said-genome-wide) | Mini MDD | 1.027 | 0.451 | 0.959 | 1.100 | 47295 |
| CRP (said-genome-wide) | Mini GAD | 1.023 | 0.359 | 0.975 | 1.073 | 47295 |
| IL-6 (ahluwalia-genome-wide) | Mini depression | 1.006 | 0.817 | 0.953 | 1.063 | 47136 |
| IL-6 (ahluwalia-genome-wide) | Mini anxiety | 1.008 | 0.674 | 0.973 | 1.044 | 47295 |
| IL-6 (ahluwalia-genome-wide) | Mini MDD | 0.992 | 0.824 | 0.926 | 1.063 | 47295 |
| IL-6 (ahluwalia-genome-wide) | Mini GAD | 1.014 | 0.560 | 0.967 | 1.064 | 47295 |

***Table 10. Standardised GRS on continuous outcomes (1st degree relatives removed)***

| Standardised GRS | Standardised Outcomes | Beta | p-value | LCI | UCI | N |
|---|---|---|---|---|---|---|
| CRP (said-cis) | Identify Task – reaction time | -0.004049836 | 0.418 | -0.013858126 | 0.005758454 | 31248 |
| CRP (said-cis) | Detect Task – reaction time | 0.007610043 | 0.137 | -0.00240993 | 0.017630016 | 30872 |
| CRP (said-cis) | OCL Task - accuracy | -0.007719335 | 0.156 | -0.018391409 | 0.002952738 | 32182 |
| CRP (said-cis) | One-back Task - accuracy | -0.003619921 | 0.506 | -0.014281494 | 0.007041651 | 31805 |
| CRP (said-cis) | RFFT | 0.005215785 | 0.321 | -0.005090252 | 0.015521822 | 32250 |
| CRP (said-cis) | PANAS – negative | 0.009218252 | 0.033 | 0.000758222 | 0.017678281 | 50275 |
| CRP (said-cis) | PANAS - positive | 0.005766673 | 0.189 | -0.002841032 | 0.014374379 | 50275 |
| GlycA (borges) | Identify Task – reaction time | 0.002430414 | 0.626 | -0.007341182 | 0.012202009 | 31248 |
| GlycA (borges) | Detect Task – reaction time | 0.004379256 | 0.390 | -0.005610349 | 0.014368861 | 30872 |
| GlycA (borges) | OCL Task - accuracy | 0.003578283 | 0.510 | -0.007055413 | 0.014211979 | 32182 |
| GlycA (borges) | One-back Task - accuracy | -0.000115487 | 0.983 | -0.010745161 | 0.010514187 | 31805 |
| GlycA (borges) | RFFT | -0.005756899 | 0.274 | -0.016072422 | 0.004558624 | 32250 |
| GlycA (borges) | PANAS – negative | 0.011174486 | 0.010 | 0.0027246 | 0.019624373 | 50275 |
| GlycA (borges) | PANAS - positive | -0.003692607 | 0.400 | -0.012290264 | 0.004905049 | 50275 |
| IL-6R (ahluwalia-cis) | Identify Task – reaction time | -0.007388214 | 0.139 | -0.017182749 | 0.00240632 | 31248 |
| IL-6R (ahluwalia-cis) | Detect Task – reaction time | -0.002237753 | 0.662 | -0.012257602 | 0.007782095 | 30872 |
| IL-6R (ahluwalia-cis) | OCL Task - accuracy | -0.005819009 | 0.285 | -0.016485395 | 0.004847377 | 32182 |
| IL-6R (ahluwalia-cis) | One-back Task - accuracy | 0.001228477 | 0.821 | -0.00942373 | 0.011880684 | 31805 |
| IL-6R (ahluwalia-cis) | RFFT | -0.007359532 | 0.162 | -0.017685313 | 0.002966249 | 32250 |
| IL-6R (ahluwalia-cis) | PANAS – negative | 0.004652218 | 0.281 | -0.003801535 | 0.01310597 | 50275 |
| IL-6R (ahluwalia-cis) | PANAS - positive | -0.003825848 | 0.383 | -0.012426959 | 0.004775262 | 50275 |
| sIL6R (rosa) | Identify Task – reaction time | -0.006482964 | 0.194 | -0.016265604 | 0.003299675 | 31248 |
| sIL6R (rosa) | Detect Task – reaction time | -0.000565435 | 0.912 | -0.01057052 | 0.00943965 | 30872 |
| sIL6R (rosa) | OCL Task - accuracy | -0.012981218 | 0.017 | -0.023632487 | -0.00232995 | 32182 |
| sIL6R (rosa) | One-back Task - accuracy | -0.005672935 | 0.296 | -0.01631509 | 0.00496922 | 31805 |
| sIL6R (rosa) | RFFT | -0.001518224 | 0.773 | -0.011827491 | 0.008791043 | 32250 |
| sIL6R (rosa) | PANAS – negative | 0.005268752 | 0.222 | -0.00318464 | 0.013722145 | 50275 |
| sIL6R (rosa) | PANAS - positive | -0.003589323 | 0.413 | -0.012190103 | 0.005011458 | 50275 |
| IL-6R (sarwar) | Identify Task – reaction time | -0.005229929 | 0.294 | -0.015001331 | 0.004541473 | 31248 |
| IL-6R (sarwar) | Detect Task – reaction time | -0.002107915 | 0.679 | -0.012098351 | 0.007882521 | 30872 |

| | | | | | | |
|---|---|---|---|---|---|---|
| IL-6R (sarwar) | OCL Task - accuracy | -0.008406874 | 0.121 | -0.019045513 | 0.002231766 | 32182 |
| IL-6R (sarwar) | One-back Task - accuracy | 0.001261055 | 0.816 | -0.009362978 | 0.011885087 | 31805 |
| IL-6R (sarwar) | RFFT | -0.00224542 | 0.670 | -0.01256145 | 0.008070611 | 32250 |
| IL-6R (sarwar) | PANAS – negative | 0.004621141 | 0.284 | -0.003829182 | 0.013071463 | 50275 |
| IL-6R (sarwar) | PANAS - positive | -0.007908285 | 0.071 | -0.016505693 | 0.000689122 | 50275 |
| CRP (said-genome-wide) | Identify Task – reaction time | -0.004384599 | 0.379 | -0.014157457 | 0.00538826 | 31248 |
| CRP (said-genome-wide) | Detect Task – reaction time | 0.005849798 | 0.251 | -0.004130549 | 0.015830146 | 30872 |
| CRP (said-genome-wide) | OCL Task - accuracy | 0.006495752 | 0.231 | -0.004134355 | 0.017125859 | 32182 |
| CRP (said-genome-wide) | One-back Task - accuracy | 0.006311614 | 0.244 | -0.00430412 | 0.016927349 | 31805 |
| CRP (said-genome-wide) | RFFT | 0.005411245 | 0.306 | -0.004946475 | 0.015768965 | 32250 |
| CRP (said-genome-wide) | PANAS – negative | -0.004457444 | 0.302 | -0.012925397 | 0.004010509 | 50275 |
| CRP (said-genome-wide) | PANAS - positive | -0.000471111 | 0.915 | -0.009086725 | 0.008144503 | 50275 |
| IL-6 (ahluwalia-genome-wide) | Identify Task – reaction time | -0.007394591 | 0.141 | -0.017231994 | 0.002442813 | 31248 |
| IL-6 (ahluwalia-genome-wide) | Detect Task – reaction time | -0.003216338 | 0.531 | -0.013280611 | 0.006847936 | 30872 |
| IL-6 (ahluwalia-genome-wide) | OCL Task - accuracy | -0.004718069 | 0.388 | -0.015431196 | 0.005995059 | 32182 |
| IL-6 (ahluwalia-genome-wide) | One-back Task - accuracy | 0.001088414 | 0.842 | -0.009610476 | 0.011787304 | 31805 |
| IL-6 (ahluwalia-genome-wide) | RFFT | -0.007272177 | 0.169 | -0.017646042 | 0.003101689 | 32250 |
| IL-6 (ahluwalia-genome-wide) | PANAS – negative | 0.004892038 | 0.259 | -0.003599313 | 0.013383388 | 50275 |
| IL-6 (ahluwalia-genome-wide) | PANAS - positive | -0.001706063 | 0.699 | -0.010345489 | 0.006933363 | 50275 |

***Table 11. Standardised GRS on continuous outcomes (2nd degree relatives removed)***

| Standardised GRS | Standardised Outcomes | Beta | p-value | LCI | UCI | N |
|---|---|---|---|---|---|---|
| CRP (said-cis) | Identify Task – reaction time | -0.004455903 | 0.375 | -0.014297241 | 0.005385435 | 30885 |
| CRP (said-cis) | Detect Task – reaction time | 0.006944798 | 0.177 | -0.003128157 | 0.017017752 | 30514 |
| CRP (said-cis) | OCL Task - accuracy | -0.006845139 | 0.211 | -0.017576303 | 0.003886025 | 31806 |
| CRP (said-cis) | One-back Task - accuracy | -0.003839799 | 0.482 | -0.014552201 | 0.006872603 | 31430 |
| CRP (said-cis) | RFFT | 0.006171204 | 0.243 | -0.004197376 | 0.016539785 | 31862 |
| CRP (said-cis) | PANAS – negative | 0.009857729 | 0.023 | 0.001341303 | 0.018374155 | 49586 |
| CRP (said-cis) | PANAS - positive | 0.005861774 | 0.184 | -0.002791759 | 0.014515308 | 49586 |
| GlycA (borges) | Identify Task – reaction time | 0.001265473 | 0.801 | -0.008551323 | 0.01108227 | 30885 |
| GlycA (borges) | Detect Task – reaction time | 0.002739723 | 0.593 | -0.007313541 | 0.012792988 | 30514 |
| GlycA (borges) | OCL Task - accuracy | 0.004528117 | 0.407 | -0.006177087 | 0.015233321 | 31806 |
| GlycA (borges) | One-back Task - accuracy | 0.001261024 | 0.817 | -0.009430819 | 0.011952867 | 31430 |
| GlycA (borges) | RFFT | -0.004472728 | 0.398 | -0.014855337 | 0.00590988 | 31862 |
| GlycA (borges) | PANAS – negative | 0.012289134 | 0.005 | 0.003775534 | 0.020802734 | 49586 |
| GlycA (borges) | PANAS - positive | -0.00339255 | 0.442 | -0.012043564 | 0.005258464 | 49586 |
| IL-6R (ahluwalia-cis) | Identify Task – reaction time | -0.006629579 | 0.187 | -0.01647197 | 0.003212811 | 30885 |
| IL-6R (ahluwalia-cis) | Detect Task – reaction time | -0.000922534 | 0.858 | -0.011008193 | 0.009163125 | 30514 |
| IL-6R (ahluwalia-cis) | OCL Task - accuracy | -0.005376679 | 0.326 | -0.016116486 | 0.005363128 | 31806 |
| IL-6R (ahluwalia-cis) | One-back Task - accuracy | 0.001617299 | 0.767 | -0.009098969 | 0.012333567 | 31430 |
| IL-6R (ahluwalia-cis) | RFFT | -0.007750897 | 0.144 | -0.018146283 | 0.002644489 | 31862 |
| IL-6R (ahluwalia-cis) | PANAS – negative | 0.004544888 | 0.296 | -0.003972594 | 0.01306237 | 49586 |
| IL-6R (ahluwalia-cis) | PANAS - positive | -0.003255724 | 0.461 | -0.011910083 | 0.005398635 | 49586 |
| sIL6R (rosa) | Identify Task – reaction time | -0.005873464 | 0.242 | -0.015704021 | 0.003957093 | 30885 |
| sIL6R (rosa) | Detect Task – reaction time | 0.000973275 | 0.850 | -0.009097362 | 0.011043913 | 30514 |
| sIL6R (rosa) | OCL Task - accuracy | -0.012447692 | 0.023 | -0.023172184 | -0.001723201 | 31806 |
| sIL6R (rosa) | One-back Task - accuracy | -0.005575441 | 0.307 | -0.016281637 | 0.005130756 | 31430 |
| sIL6R (rosa) | RFFT | -0.002065713 | 0.697 | -0.012447815 | 0.008316389 | 31862 |
| sIL6R (rosa) | PANAS – negative | 0.005671932 | 0.192 | -0.00284578 | 0.014189644 | 49586 |
| sIL6R (rosa) | PANAS - positive | -0.003427152 | 0.438 | -0.012081793 | 0.00522749 | 49586 |
| IL-6R (sarwar) | Identify Task – reaction time | -0.004503529 | 0.369 | -0.014321039 | 0.005313981 | 30885 |
| IL-6R (sarwar) | Detect Task – reaction time | -0.000572008 | 0.911 | -0.010626347 | 0.009482331 | 30514 |

| | | | | | | |
|---|---|---|---|---|---|---|
| IL-6R (sarwar) | OCL Task - accuracy | -0.007931195 | 0.147 | -0.018641834 | 0.002779444 | 31806 |
| IL-6R (sarwar) | One-back Task - accuracy | 0.001561761 | 0.775 | -0.00912495 | 0.012248472 | 31430 |
| IL-6R (sarwar) | RFFT | -0.002749847 | 0.604 | -0.01313688 | 0.007637186 | 31862 |
| IL-6R (sarwar) | PANAS – negative | 0.004408061 | 0.310 | -0.004106041 | 0.012922162 | 49586 |
| IL-6R (sarwar) | PANAS - positive | -0.007323901 | 0.097 | -0.015974627 | 0.001326825 | 49586 |
| CRP (said-genome-wide) | Identify Task – reaction time | -0.00441728 | 0.377 | -0.014226265 | 0.005391706 | 30885 |
| CRP (said-genome-wide) | Detect Task – reaction time | 0.006311434 | 0.218 | -0.003722837 | 0.016345706 | 30514 |
| CRP (said-genome-wide) | OCL Task - accuracy | 0.006649428 | 0.223 | -0.004041917 | 0.017340774 | 31806 |
| CRP (said-genome-wide) | One-back Task - accuracy | 0.005929476 | 0.276 | -0.004738813 | 0.016597765 | 31430 |
| CRP (said-genome-wide) | RFFT | 0.004949002 | 0.352 | -0.005479586 | 0.01537759 | 31862 |
| CRP (said-genome-wide) | PANAS – negative | -0.004004421 | 0.358 | -0.012537485 | 0.004528644 | 49586 |
| CRP (said-genome-wide) | PANAS - positive | -0.000744121 | 0.866 | -0.009414337 | 0.007926095 | 49586 |
| IL-6 (ahluwalia-genome-wide) | Identify Task – reaction time | -0.006600991 | 0.191 | -0.016486541 | 0.00328456 | 30885 |
| IL-6 (ahluwalia-genome-wide) | Detect Task – reaction time | -0.001896891 | 0.714 | -0.012027463 | 0.008233682 | 30514 |
| IL-6 (ahluwalia-genome-wide) | OCL Task - accuracy | -0.004187567 | 0.447 | -0.014974478 | 0.006599344 | 31806 |
| IL-6 (ahluwalia-genome-wide) | One-back Task - accuracy | 0.001494971 | 0.785 | -0.009268274 | 0.012258216 | 31430 |
| IL-6 (ahluwalia-genome-wide) | RFFT | -0.007700978 | 0.148 | -0.018144867 | 0.002742911 | 31862 |
| IL-6 (ahluwalia-genome-wide) | PANAS – negative | 0.004759201 | 0.276 | -0.0037966 | 0.013315002 | 49586 |
| IL-6 (ahluwalia-genome-wide) | PANAS - positive | -0.001113178 | 0.802 | -0.009806523 | 0.007580167 | 49586 |

***Table 12. Standardised GRS on continuous outcomes (3rd degree relatives removed)***

| Standardised GRS | Standardised Outcomes | Beta | p-value | LCI | UCI | N |
|---|---|---|---|---|---|---|
| CRP (said-cis) | Identify Task – reaction time | -0.002774482 | 0.586 | -0.012749699 | 0.007200734 | 30055 |
| CRP (said-cis) | Detect Task – reaction time | 0.007615498 | 0.143 | -0.002576949 | 0.017807945 | 29688 |
| CRP (said-cis) | OCL Task - accuracy | -0.007644057 | 0.168 | -0.018519803 | 0.003231689 | 30948 |
| CRP (said-cis) | One-back Task - accuracy | -0.003689577 | 0.506 | -0.014557446 | 0.007178292 | 30581 |
| CRP (said-cis) | RFFT | 0.006567816 | 0.220 | -0.003937313 | 0.017072945 | 30978 |
| CRP (said-cis) | PANAS – negative | 0.010495546 | 0.017 | 0.001849709 | 0.019141384 | 48230 |
| CRP (said-cis) | PANAS - positive | 0.005831615 | 0.193 | -0.002940174 | 0.014603405 | 48230 |
| GlycA (borges) | Identify Task – reaction time | -0.000180337 | 0.972 | -0.010122833 | 0.009762159 | 30055 |
| GlycA (borges) | Detect Task – reaction time | 0.002597782 | 0.617 | -0.007573292 | 0.012768857 | 29688 |
| GlycA (borges) | OCL Task - accuracy | 0.004564767 | 0.409 | -0.006281792 | 0.015411325 | 30948 |
| GlycA (borges) | One-back Task - accuracy | 0.000830935 | 0.881 | -0.010012673 | 0.011674543 | 30581 |
| GlycA (borges) | RFFT | -0.005200444 | 0.332 | -0.015716549 | 0.00531566 | 30978 |
| GlycA (borges) | PANAS – negative | 0.010745678 | 0.015 | 0.00209993 | 0.019391427 | 48230 |
| GlycA (borges) | PANAS - positive | -0.003318834 | 0.458 | -0.012090663 | 0.005452995 | 48230 |
| IL-6R (ahluwalia-cis) | Identify Task – reaction time | -0.00691165 | 0.174 | -0.016878898 | 0.003055598 | 30055 |
| IL-6R (ahluwalia-cis) | Detect Task – reaction time | -0.000272639 | 0.958 | -0.010472643 | 0.009927365 | 29688 |
| IL-6R (ahluwalia-cis) | OCL Task - accuracy | -0.004406893 | 0.427 | -0.01528395 | 0.006470165 | 30948 |
| IL-6R (ahluwalia-cis) | One-back Task - accuracy | 0.001232496 | 0.824 | -0.009631128 | 0.012096121 | 30581 |
| IL-6R (ahluwalia-cis) | RFFT | -0.006775222 | 0.207 | -0.017307765 | 0.003757322 | 30978 |
| IL-6R (ahluwalia-cis) | PANAS – negative | 0.005925359 | 0.179 | -0.002721924 | 0.014572643 | 48230 |
| IL-6R (ahluwalia-cis) | PANAS - positive | -0.003399121 | 0.448 | -0.012172129 | 0.005373886 | 48230 |
| sIL6R (rosa) | Identify Task – reaction time | -0.006150564 | 0.226 | -0.016099049 | 0.00379792 | 30055 |
| sIL6R (rosa) | Detect Task – reaction time | 0.001400574 | 0.787 | -0.008776395 | 0.011577544 | 29688 |
| sIL6R (rosa) | OCL Task - accuracy | -0.011708759 | 0.035 | -0.022564667 | -0.000852851 | 30948 |
| sIL6R (rosa) | One-back Task - accuracy | -0.00587703 | 0.288 | -0.016724268 | 0.004970208 | 30581 |
| sIL6R (rosa) | RFFT | -0.000927542 | 0.863 | -0.011439615 | 0.009584532 | 30978 |
| sIL6R (rosa) | PANAS – negative | 0.006161837 | 0.162 | -0.002482684 | 0.014806357 | 48230 |
| sIL6R (rosa) | PANAS - positive | -0.003788907 | 0.397 | -0.012559112 | 0.004981298 | 48230 |
| IL-6R (sarwar) | Identify Task – reaction time | -0.005075206 | 0.317 | -0.01501438 | 0.004863967 | 30055 |
| IL-6R (sarwar) | Detect Task – reaction time | -0.000128678 | 0.980 | -0.010293975 | 0.010036619 | 29688 |

| | | | | | | |
|---|---|---|---|---|---|---|
| IL-6R (sarwar) | OCL Task - accuracy | -0.005936317 | 0.283 | -0.016781671 | 0.004909036 | 30948 |
| IL-6R (sarwar) | One-back Task - accuracy | 0.001771228 | 0.749 | -0.009060039 | 0.012602496 | 30581 |
| IL-6R (sarwar) | RFFT | -0.00101124 | 0.851 | -0.011533065 | 0.009510584 | 30978 |
| IL-6R (sarwar) | PANAS – negative | 0.005588038 | 0.205 | -0.00305536 | 0.014231436 | 48230 |
| IL-6R (sarwar) | PANAS - positive | -0.007725396 | 0.084 | -0.016494226 | 0.001043433 | 48230 |
| CRP (said-genome-wide) | Identify Task – reaction time | -0.003617212 | 0.475 | -0.013545849 | 0.006311426 | 30055 |
| CRP (said-genome-wide) | Detect Task – reaction time | 0.006502643 | 0.209 | -0.003633692 | 0.016638977 | 29688 |
| CRP (said-genome-wide) | OCL Task - accuracy | 0.005456883 | 0.323 | -0.005364119 | 0.016277884 | 30948 |
| CRP (said-genome-wide) | One-back Task - accuracy | 0.004386723 | 0.426 | -0.00642168 | 0.015195127 | 30581 |
| CRP (said-genome-wide) | RFFT | 0.004065849 | 0.450 | -0.006483806 | 0.014615503 | 30978 |
| CRP (said-genome-wide) | PANAS – negative | -0.002859669 | 0.517 | -0.011515421 | 0.005796083 | 48230 |
| CRP (said-genome-wide) | PANAS - positive | -0.000701674 | 0.876 | -0.009483198 | 0.00807985 | 48230 |
| IL-6 (ahluwalia-genome-wide) | Identify Task – reaction time | -0.006858244 | 0.179 | -0.016872277 | 0.00315579 | 30055 |
| IL-6 (ahluwalia-genome-wide) | Detect Task – reaction time | -0.001390068 | 0.790 | -0.011638639 | 0.008858503 | 29688 |
| IL-6 (ahluwalia-genome-wide) | OCL Task - accuracy | -0.003063095 | 0.583 | -0.013990921 | 0.00786473 | 30948 |
| IL-6 (ahluwalia-genome-wide) | One-back Task - accuracy | 0.001092963 | 0.844 | -0.009821351 | 0.012007278 | 30581 |
| IL-6 (ahluwalia-genome-wide) | RFFT | -0.006651944 | 0.218 | -0.017237636 | 0.003933748 | 30978 |
| IL-6 (ahluwalia-genome-wide) | PANAS – negative | 0.006212709 | 0.161 | -0.002475698 | 0.014901116 | 48230 |
| IL-6 (ahluwalia-genome-wide) | PANAS - positive | -0.001142555 | 0.799 | -0.009957346 | 0.007672235 | 48230 |

#### SUPPLEMENTARY FIGURES

***Figure 1. Flow diagram of total sample size available for genetic risk score analysis.***

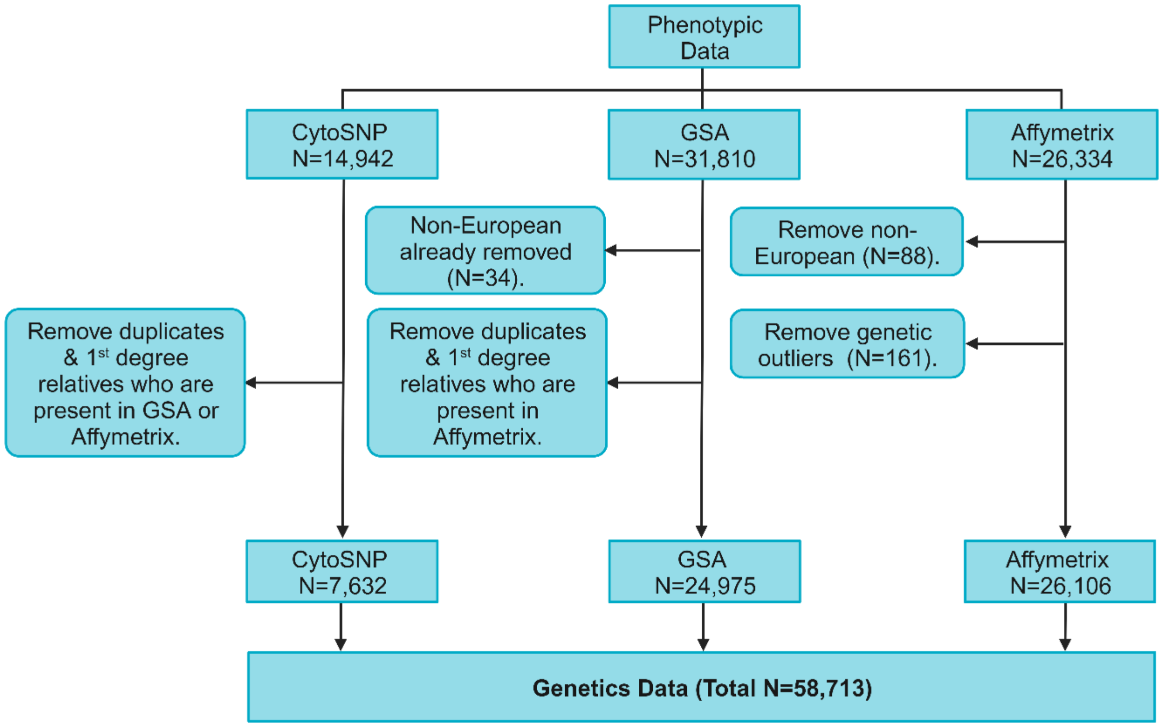

*Note:* Phenotypic data includes individuals in Lifelines who are ≥ 18 years and excludes individuals who self-report one of the following health conditions: Alzheimer's disease, dementia, epilepsy, multiple sclerosis, Parkinson's disease, stroke. Duplicates and first-degree relatives were removed from CytoSNP where possible due to its poorer imputation quality in comparison to the other chips. Lists of duplicates and first-degree relatives to remove between chips were provided by Lifelines. Genetic outliers and non-European individuals were also provided by Lifelines. Figure created using *Biorender.com*.
